# Enhancing automated indexing of publication types and study designs in biomedical literature using full-text features

**DOI:** 10.1101/2025.04.23.25326300

**Authors:** Joe D. Menke, Shufan Ming, Shruthan Radhakrishna, Halil Kilicoglu, Neil R. Smalheiser

**Affiliations:** School of Information Sciences, University of Illinois Urbana-Champaign, 501 E Daniel Street, Champaign, 61820, IL, USA; Department of Computer Science, University of Illinois Urbana-Champaign, 201 North Goodwin Avenue, Urbana, 61801, IL, USA; Department of Psychiatry, University of Illinois Chicago, 1601 W Taylor Street, Chicago, 60612, IL, USA

**Keywords:** natural language processing, literature mining, publication type indexing, study design indexing, information retrieval, evidence synthesis

## Abstract

**Objective:** Searching for biomedical articles by publication type or study design is essential for tasks like evidence synthesis. Prior work has relied solely on PubMed information or addressed a limited set of types (e.g., randomized controlled trials). In this study, we build on previous work by leveraging full-text features, enriched text representations, and advanced optimization techniques for comprehensive indexing.

**Methods:** Using a dataset of PubMed articles published between 1987 and 2023 with human-annotated indexing terms, we fine-tuned BERT-based encoders (PubMedBERT, BioLinkBERT, SPECTER, SPECTER2-Base, SPECTER2-Clf) to investigate whether text representations based on different pre-training objectives could benefit the task. We incorporated textual and verbalized metadata features, full-text extraction (rule-based, extractive, and abstractive summarization), and additional topical information about the articles. To mitigate potential label noise and improve calibration, we used asymmetric loss and label smoothing. We also explored contrastive learning approaches (SimCSE, ADNCE, HeroCon, WeighCon). Models were evaluated using precision, recall, F1 score (both micro- and macro-), and area under ROC curve (AUC).

**Results:** Fine-tuning SPECTER2-Base with asymmetric loss, label smoothing and contrastive learning (ADNCE and HeroCon) improved performance significantly over the previous best model (micro-F1: 0.658 → 0.670 [+1.8%]; macro-F1: 0.643 → 0.677 [+5.3%]; p < 0.001). Asymmetric loss and using SPECTER2-Base instead of PubMedBERT contributed most to this gain, while contrastive learning provided more moderate gains. Full-text features boosted performance by 2.4% (micro-F1) and 0.8% (macro-F1) over the baseline (micro-F1: 0.656 → 0.672; macro-F1: 0.595 → 0.600; p < 0.001).

**Conclusion:** Full-text features, citation-aware encoders, and fine-tuning optimizations significantly improve publication type and study design indexing. Future work should refine label accuracy, better distill relevant full-text information, and expand label sets to meet needs of the research community. Data, code, and models are available at https://github.com/ScienceNLP-Lab/MultiTagger-v2.

**Highlights:**

- We trained and validated Transformer-based models for automatic indexing of publication types and study designs in biomedical articles, using a dataset with 61 labels derived primarily from expert-assigned PubMed indexing terms.
- We investigated whether enriched article representations, advanced optimization techniques, and fine-grained labels could enhance model performance.
- The largest performance improvement came from using citation-aware article representations and asymmetric loss.
- Models trained using full-text features outperformed models trained using PubMed-only features, demonstrating the utility of full-text content for this task.

**Graphical Abstract:** 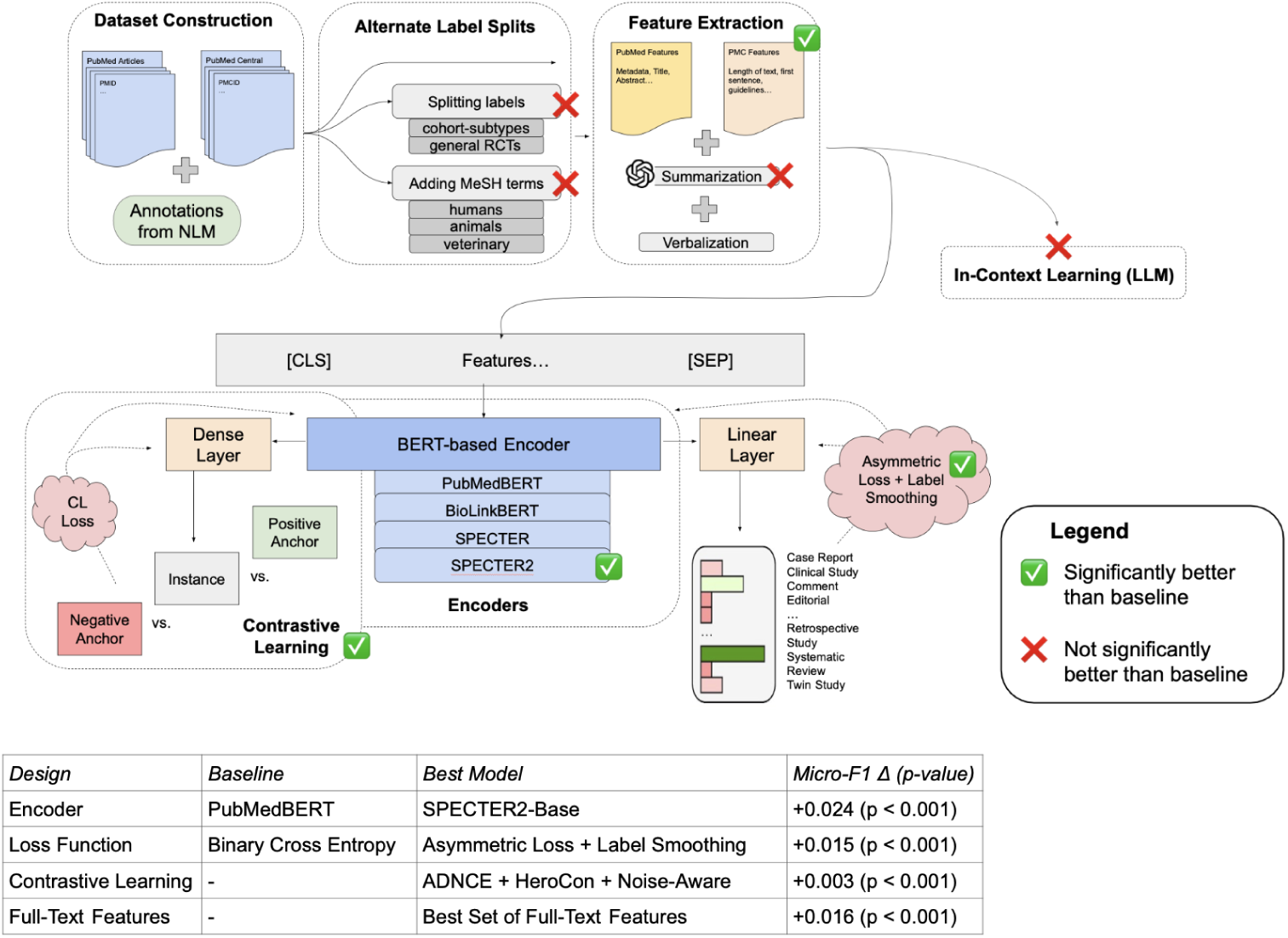

## 1. Introduction

The ability to search and filter research publications efficiently is crucial for all researchers. This is especially true in areas such as evidence-based medicine, which uses the best available clinical evidence to inform treatment [1]. The size and rapid expansion of biomedical literature poses a significant challenge, making the identification of relevant publications a lengthy and labor-intensive task [2, 3, 4]. Improvements to literature screening processes are poised to significantly benefit downstream tasks, including evidence synthesis.

The U.S. National Library of Medicine (NLM) indexes publications in MEDLINE, their bibliographic database, by Medical Subject Headings (MeSH) and publication types (PT) to help facilitate article retrieval. Historically, the task of indexing MEDLINE articles has been carried out manually by skilled medical indexing experts [5]. Over the past two decades, NLM’s Medical Text Indexer (MTI) program has increasingly automated article indexing [6, 7, 8, 9]. The most recent MTI model uses a combination of convolutional neural networks (CNN) and PubMedBERT to generate and rank MeSH candidates [9]. Since 2022, automatic MeSH indexing has been applied to all journals indexed for MEDLINE with human indexers performing subsequent review and curation to ensure accuracy and completeness, which reduced the average indexing time from 145 days down to just one day [10].

Outside of the NLM, the challenge of automatically indexing the biomedical literature continues to garner substantial attention. For example, since 2013, the annual BioASQ shared tasks have focused on advancing MeSH indexing methods with MTI often serving as a baseline [11, 12, 13]. Notable approaches include ensembling models using a learning-to-rank framework (e.g., MeSHLabeler[14])), combining learning-to-rank with deep representations (e.g., DeepMESH [15]), using contextualized representations and fine-tuning (e.g., BERTMeSH [16]), incorporating full-text content (FullMeSH [17]), and models that also incorporate novel attention-based mechanisms (e.g., MeSHProbeNet-P [18], KenMeSH [19]). The focus of these methods is to assign MeSH headings and subheadings to publications. These headings are primarily based on an article’s topic of study, e.g., “Brain” or “Asthma”. However, this is not the case for all MeSH terms. A relatively small subset of MeSH terms focus on methodological characteristics of research, indicating how a study was conducted rather than its subject matter, e.g., Cohort Studies or Double-Blind Method. This information is particularly important for evidence synthesis pipelines, serving as an initial automated filtering step before manual analysis and synthesis [20, 21, 22, 23].

A significant body of work has emerged focusing on these study design-related terms as well as publication types (collectively referred to as PTs here). For example, RCT Tagger classifies Randomized Controlled Trials (RCT) using a SVM model with n-gram based features and manually annotated features from MEDLINE (i.e., MeSH terms and publication type) to support systematic reviews [24]. Building on this work, Wallace et al. [25] used similar machine learning methods along with crowdsourcing, while Marshall et al. [26] built binary RCT classifiers using ensembles of SVMs, CNNs, and MEDLINE publication type tags. Extending beyond RCTs, MultiTagger, a suite of binary SVM classifiers, was designed [27] and developed [28] to generate probabilistic estimates of 50 different PTs. Both RCT Tagger and MultiTagger demonstrated high recall in identifying RCTs, resulting in little information loss [29] at much faster speeds [30] demonstrating their value for systematic reviews. Focusing more on pre-clinical animal research, Neves et al. developed models to identify study designs that may serve as alternatives to animal experiments (e.g., *in vitro*) [31]. Most recently, we developed a new version of MultiTagger [32] formulating the task as multi-label classification and fine-tuning PubMedBERT [33] using text (title, abstract) and metadata features from PubMed to improve micro-F1 by 40% (0.497 *→* 0.697) and macro-F1 by 52% (0.416 *→* 0.632) compared to the original MultiTagger [28]. This comparison focused on 49 labels tagged by the original MultiTagger and was performed using a test set of 64,400 PubMed articles with titles and abstracts longer than 25 characters.

In this study, we extend our prior work in several directions and make the following contributions:

- We investigate whether (and which) full-text features could improve PT classification. In prior work, full-text information was shown to benefit MeSH indexing [16, 17]. Due to the context size limitations of BERT-based models (512 tokens), which is much smaller than the average full-text article, and based on the hypothesis that most full-text content is irrelevant to PT classification, we conduct experiments with extractive and abstractive summarization methods.
- We assess whether enriched document representations could benefit PT classification task. Specifically, we hypothesize that publications may cite other studies that share methodological similarities and fine-tune several Transformer-based models that are pre-trained in part using citation-related information (BioLinkBERT [34] and SPECTER [35]). Further, we hypothesize that, compared to PubMedBERT which uses standard masked language modeling and next sentence prediction as pre-training objectives, models pre-trained on document classification tasks could provide enriched representations that benefit the PT classification task and experiment with such models (SPECTER2 [36]). We also explore unsupervised and supervised contrastive learning in depth in an effort to better align document representations with the labels. Specifically, we compare unsupervised contrastive loss approaches (SimCSE [37] and ADjusted InfoNCE (ADNCE) [38]) with supervised approaches (HeroCon [39] and WeighCon [40]).
- We incorporate advanced optimization techniques in model fine-tuning and label splitting for mitigating the effect of potential noise and inconsistencies in PT indexing [24, 29]. Furthermore, given the hierarchical nature of some PT labels and their heterogeneity, we add labels for more fine-grained categories during training to evaluate whether more discriminative representations can be learned.
- We audit labels for a small, stratified sample of articles to assess the extent of label noise in the dataset.

|  |  |
| --- | --- |
| **Problem** | Manually indexing research articles by publication type and study design is difficult and time-consuming. |
| **What is Already Known** | Automated approaches to publication type and study design indexing focus on titles and abstracts and are mostly limited to a small number of types or use potentially noisy datasets as ground truth without consideration of this noise. |
| **What This Paper Adds** | We enhance publication type and study design indexing using full-text features, enriched article representations, and noise-aware training techniques. |
| **Who Would Benefit** | Researchers and clinicians who use research repositories (e.g., PubMed), especially those who use publication types and study designs for article screening and evidence synthesis. |

## 2. Materials and Methods

The components of our overall approach and the processing pipeline is illustrated in Figure 1. We describe each component below. We also discuss the experiments we performed and the evaluation methodology at the end of this section.

**Figure 1:**
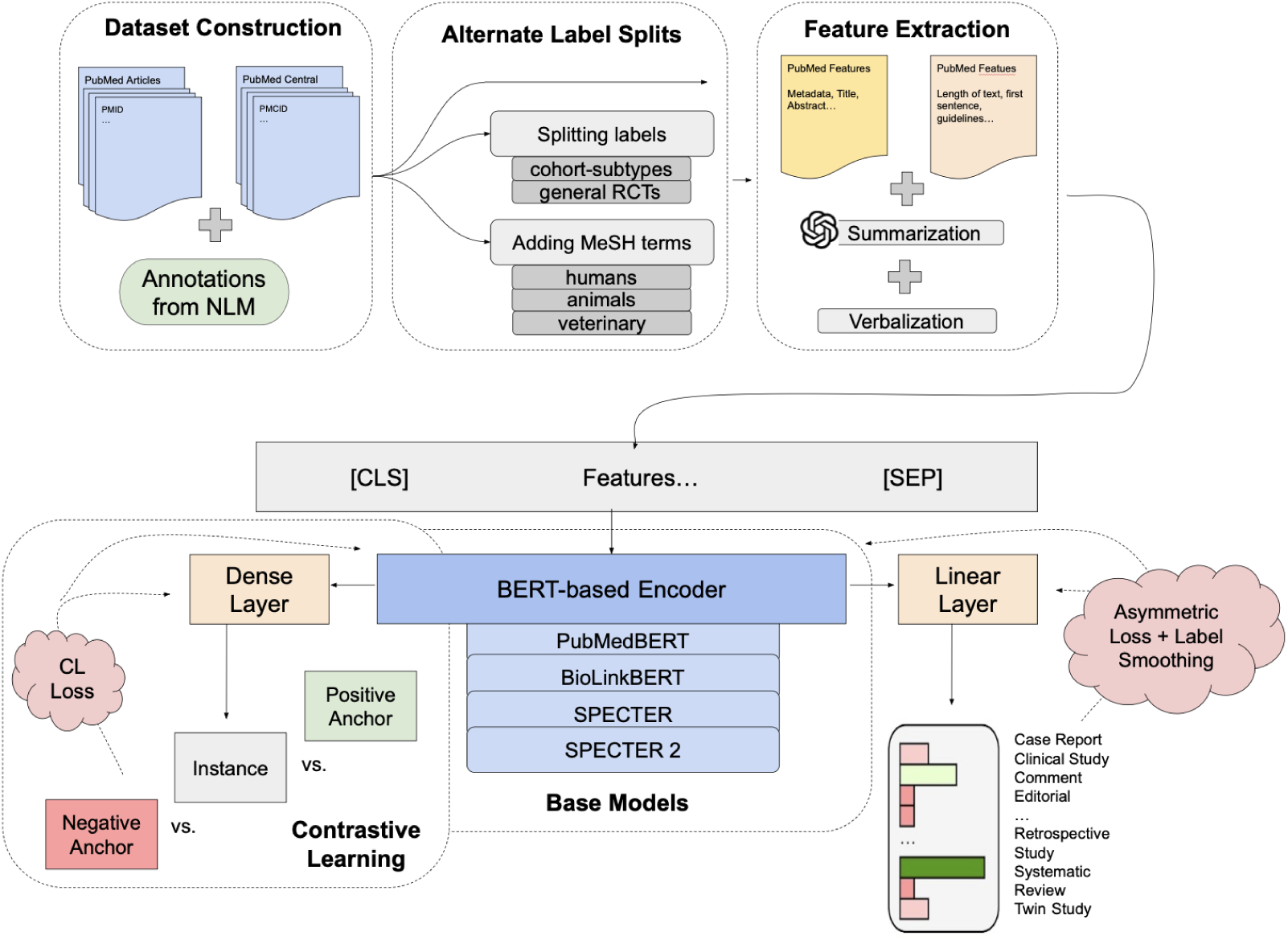
Flow diagram of dataset and feature construction as well as experiments including feature extraction, alternate label splits, base encoders, and contrastive learning.

### 2.1. Dataset Construction

Following prior work [9], we included articles with PT labels that were manually assigned by trained human indexers at the U.S. National Library of Medicine (NLM) based on detailed content review of the article. We excluded articles that were automatically tagged by MTI [6, 7, 8, 9] or human-curated (where curators edited or validated model-suggested labels) to improve label quality (i.e., “NOT (indexingmethod curated OR indexing-method automated)”). We also restricted the dataset to articles in English or containing an English abstract, and published between 1987 and 2023. We made several modifications to PT labels to further improve label quality and consistency:

- For Systematic Review and Systematic Reviews as Topic, we combined keyword searches with manually assigned labels. These publication types were introduced in 2019, and automatic retrospective indexing of earlier articles using heuristic methods led to label noise. [28].
- Veterinary Clinical Trials and Veterinary RCTs also used the keyword “human” to exclude articles involving human research (i.e., “NOT humans[ti]”), as we found that manual NLM annotations for veterinary PTs were sparse and unreliable even for the articles published after the introduction of these PTs in 2019.
- Relatedly, for human RCTs, we require the “Humans” MeSH term to better differentiate between human RCTs and veterinary RCTs.
- For a subset of clinical PTs (e.g., RCTs, Cohort Studies), we excluded articles also labeled as Editorial, Letter, Comment, Practice Guideline, or Review, as these were commonly found to simply discuss the PT rather than be of that type, following previous work [28, 32]. For example, if an article is tagged as RCT and Editorial, we only consider it Editorial.
- Retraction of Publication included articles between 1987 and 2025 to increase the number of positively labeled articles.
- Scientific Integrity Review utilized all NLM labels (human-labeled, human-curated, or automated) as this PT has easily discernible features (i.e., title is generally “Findings of Research Misconduct”) where including human-curated or automatically-tagged articles did not appear to lower label quality.

Except systematic review- and veterinary-related labels above, we did not generate, modify, or filter any other PT labels using textual patterns or keyword-based heuristics.

The searches used for dataset construction for all PTs are available at the project GitHub repository. As an example, we used the following search to retrieve the articles with the Clinical Trial PT:

> (1987:2023[dp] AND (english[Language] OR english abstract[pt]) NOT (indexingmethod curated OR indexingmethod automated)) AND “clinical trial”[pt] NOT case-control studies[mh:noexp] NOT cohort studies[mh:noexp] NOT editorial[pt] NOT letter[pt] NOT comment[pt] NOT “practice guideline”[pt] NOT review[pt]

In total, we retrieved 9.6 million PubMed articles with 61 distinct PT labels. We sampled from this initial candidate set using a modified version of stratified sampling to restrict the dataset to a more reasonable size for training, while making efforts to prevent any majority class from dominating the dataset and retaining enough rare labels to effectively train the models. The stratified dataset was supplemented with randomly selected negative articles, i.e., articles containing no positive labels, totaling 20% of the entire dataset. For full-text experiments, we used the subset of articles in our dataset that are available in the PMC Open Access Subset.

We used PubMed identifiers (PMIDs) as unique article identifiers. Although PMIDs are generally stable, rare instances of duplicate PMIDs occur (e.g., preprints and published versions, or articles published in multiple journals). To prevent data leakage, we excluded preprints and applied two-stage deduplication. In the first stage, MinHash locality-sensitive hashing (LSH) was used to identify candidate duplicate article pairs where the Jaccard similarity exceeded 0.7. In the second stage, these candidate pairs were verified using the Levenshtein similarity ratio, with duplicate articles defined as those exceeding 0.85 similarity for concatenated title–abstract–publication date strings and having at least 30 characters. The 0.85 cutoff was determined after manually examining articles with Levenshtein ratio greater than 0.80.

While these expert-assigned annotations have been widely used in prior work on biomedical article classification and downstream tasks such as systematic reviews [11, 13, 19, 9, 28], they may contain inconsistencies [29, 41] arising from inter-annotator variability, evolving indexing conventions, the inherent complexity of multi-label annotation, and occasional ambiguity in whether an article uses a particular study design or merely discusses it (e.g., describing double-blinding in practice). Accordingly, in this work, we treat these labels as silver standard rather than gold standard.

#### 2.1.1. Alternate Label Splits

Some PT labels are hierarchical in nature and have high topical heterogeneity, making it difficult to learn effective representations with models. In a set of experiments, we aimed to evaluate whether adding more information through more fine-grained labels could improve model performance by allowing it to better learn these heterogeneous labels. For these experiments, we added new labels into the dataset during the training process.

First, we added labels that represented overlaps between two heterogeneous PTs to steer the model to pay more attention to the correlation between these PTs. For example, Longitudinal Studies is a PT under Cohort Studies in MeSH, so a Longitudinal-Cohort Studies label was added. Positively labeled instances in this case are articles that were already labeled as both Longitudinal Studies and Cohort Studies. This was done for all children PTs of Cohort Studies (i.e., Follow-Up Studies, Longitudinal Studies, Prospective Studies, and Retrospective Studies). This experiment is referred to as cohort-split. Similarly, for the generalized rct experiment, we added a new label encompassing articles labeled as either Human RCTs or Veterinary RCTs to see if this could help the model better understand the study design (RCT), which is independent of species.

We also performed experiments distinguishing human, animal, and veterinary studies. For human and animal experiments, we added a binary label to the multi-label dataset based on the presence of specific MeSH terms for the article (“Humans”, “Animals”). We labeled articles during the veterinary experiment if they had one of the following MeSH terms: “Dogs”, “Cats”, “Cattle”, “Horses”, and “Swine”. These terms were identified based on their frequency (*>*20) in articles tagged with the Randomized Controlled Trial, Veterinary PT. Lastly, we conducted a combined experiment, where we added labels from all the following experiments during training: cohort-split, generalized rct, human, animal, and veterinary.

Note that these terms were only used during training (and not in evaluation) to ensure that the results are comparable to those of the models trained with the standard labels.

### 2.2. Feature Extraction

#### 2.2.1. PubMed Features

We mainly followed our earlier work [32] for extracting PubMed features. For each article, features (title, abstract, and other metadata) were extracted from PubMed and verbalized as model input. Feature verbalization is meant to better contextualize features in efforts to improve BERT-based representations, which are trained to rely on context within their masked language modeling task. Features and verbalizations are described in Table 1. We made the following changes to features from previous work:

- We limited acronym extraction to article titles only to capture study synonyms.
- We removed publication date as a feature.
- We added a feature related to the number of affiliations in an article.
- We combined some features to reduce input length (e.g., number of authors and affiliations).
- We verbalized numbers in metadata features (“There are 4 authors” → “There are four authors”) to prevent information loss during tokenization.

**Table 1:**
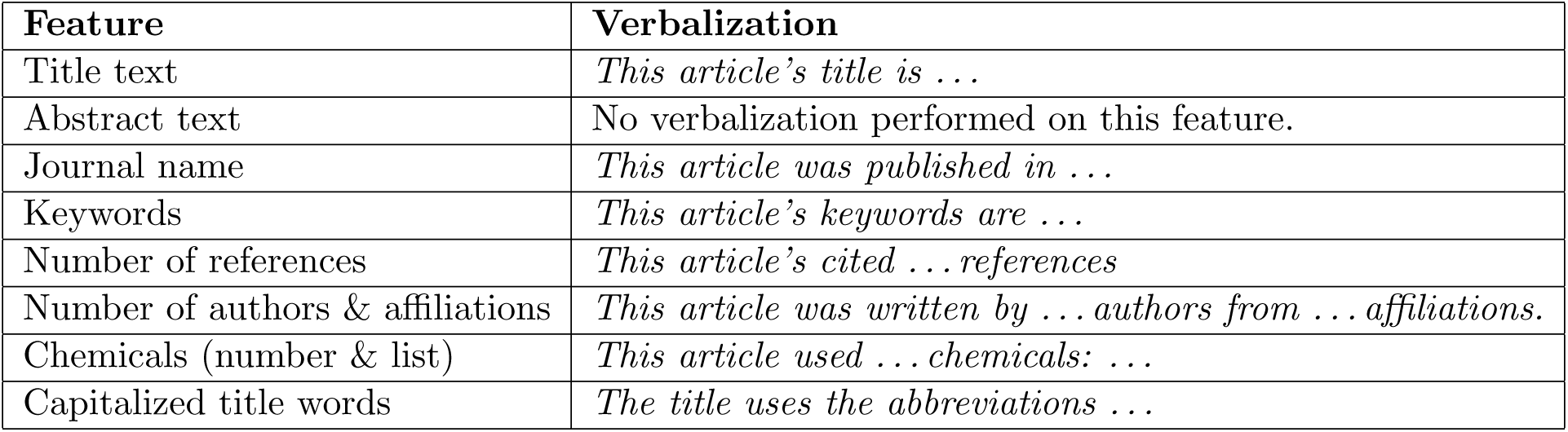
The features extracted from PubMed that were used in experiments. In cases where a feature is missing, an empty string replaces it.

| Feature | Verbalization |
| --- | --- |
| Title text | *This article’s title is ...* |
| Abstract text | No verbalization performed on this feature. |
| Journal name | *This article was published in ...* |
| Keywords | *This article’s keywords are ...* |
| Number of references | *This article’s cited ... references* |
| Number of authors & affiliations | *This article was written by ... authors from ... affiliations.* |
| Chemicals (number & list) | *This article used ... chemicals: ...* |
| Capitalized title words | *The title uses the abbreviations ...* |

#### 2.2.2. Full-Text Features

For experiments involving full-text articles, a variety of features were extracted from PubMed Central (PMC) when available. The extracted features and examples of how they were verbalized are shown in Table 2. A subset of the features are explained below, others are self-explanatory:

- *Number features* refers to features related to (1) the number of figures detected, (2) the number of tables detected, and (3) the approximate article word count.
- *Guidelines* refers to a regular expression-based feature that captures the mentions of various reporting guidelines (e.g., “CONSORT” for RCTs and “STROBE” for observational studies). Guideline mentions may provide a clue to the article PT.
- *Ethics* refers to a regular expression-based feature that captures the mentions of ethical approval. This feature could help models differentiate between PTs that commonly use humans versus those that do not.
- *Identifiers* refers to a regular expression-based rule that determines the location and frequency of clinical identifiers (e.g., NCT# from clinical-trials.gov detected in the methods section or a table).
- *1st sentence* refers to the first sentence in the Methods section (if detected). Otherwise, the first sentence in the full-text was used.
- *1st paragraph* refers to the first paragraph in the Methods section (if detected). Otherwise, the first paragraph in the full-text was used.
- *Label sentences* refers to sentences that mention any PT label (e.g., Randomized Controlled Trials), which are identified using regular expressions.

**Table 2:**
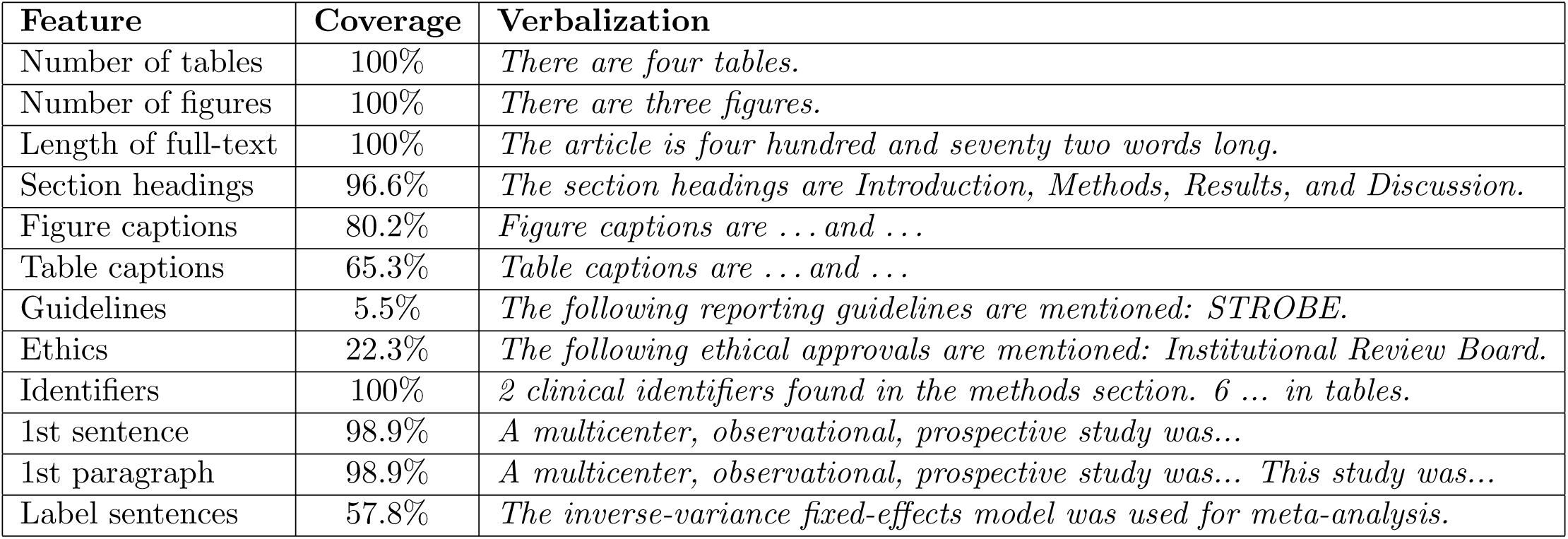
The full-text features used in full-text specific experiments, as well as examples showing how they were verbalized. Coverage represents the proportion of articles with a full-text feature extracted. Many number features are 100% as they defaulted to 0 if a table was not found.

A few features verbalized missing features: caption features (“No figure/table detected.”), guidelines (“No reporting guidelines were detected.”), ethics (“No ethical approvals detected.”), and label sentences (“No sentences containing labels were detected.”). All other features verbalized missing features using an empty string.

#### 2.2.3. Summarization Features

We also explored summarizing relevant information from full-text as input to the model. Instead of processing the entire document, we focused on sentences from the Introduction and Methods sections only (referred to as “full-text” below for brevity). This approach is motivated by two considerations. First, the model’s token limitations make it impractical to process lengthy full-text documents efficiently. Second, narrowing the input to these targeted sections provides the model with more relevant information. Our preliminary analysis of 50 articles showed that sentences containing key PT information often appear in the Introduction or Methods sections. This is important, as models often struggle with long-context memorization in summarization, which can result in hallucinations or the generation of irrelevant content [42].

We employed abstractive and extractive summarization methods to extract study design-related information from full-text. We augmented the PubMed and full-text features with summarization-based features to fine-tune the PT classification model (described below). Abstractive summarization provides a high-level overview of an entire article, often involving rewriting, paraphrasing, and reorganizing source document—processes that are prone to errors [43]. In contrast, extractive summarization ensures factual consistency by directly selecting relevant content from the source document.

##### Extractive summarization approaches

As an extractive summarization baseline, we used TextRank[44], an unsupervised graph-based ranking algorithm. It represents text as a graph where sentences are nodes, and edges represent their similarity (i.e., content overlap). By applying the PageRank algorithm, TextRank identifies the most “important” sentences based on their centrality in the graph.

We also utilized an off-the-shelf dense retriever, BMRetriever, to extract the most reliable grounding information, serving as an extractive summary [45]. This model leverages the capabilities of autoregressive large language models (LLMs) and is further fine-tuned on a combination of five biomedical tasks with labeled synthetic user query and response pairs. Given a task description, the model returns scores for each input sentence, indicating their informativeness and relevance. We experimented with three ex-tractive prompts: extractive (single), extractive (multiple), and extractive (multiple+definition). The extractive (single) experiment used a consolidated query listing all target publication type labels with the prompt:

> “Given the following categories of interest and a biomedical article, retrieve sentences that are indicative of any of these study designs or publication types.”

When all label names are presented together, the model may struggle to distinguish between the nuances of each label, especially if they are semantically similar or overlapping. This could result in retrieving sentences that are only somewhat related or not specific enough to each label. Therefore, we also employ the following multiple retrieval approaches to test our hypothesis. The extractive (multiple) experiment employed each category label as a distinct query. This approach assumes that the model understands the label’s meaning and can retrieve sentences that are semantically similar to the full-text. Finally, extractive (multiple+definition) paired category labels with brief definitions to provide additional context and enhance the retrieval process. In both methods, sentences were ranked based on their similarity scores, and the top 20 sentences were selected for further use. 20 sentences were chosen to balance the context length limitations of the model (i.e., 512 tokens) with the need to ensure sufficient coverage of potentially relevant information.

##### Abstractive summarization approaches

Longformer-Encoder-Decoder [46], a sequence-to-sequence model, has been widely studied for the abstractive summarization of long documents, including biomedical articles [47]. PRIMERA [48] is based on Longformer-Encoder-Decoder and has been further trained with a gap sentence generation objective, where salient sentences were masked to encourage the model to generate them. This model also leverages global attention and a specially pretrained doc-sep token that marks section boundaries. Together, these mechanisms allow it to aggregate information across the entire document more effectively, which may enhance memory retention and reduce hallucinations. This can be especially important in identifying study design information, which can be implicit and may need to be inferred across sentences or sections.

Additionally, we prompted a large language model (meta-llama/Llama-3.2-3B-Instruct [49]) to generate an open-ended, publication-type-focused summary as auxiliary input, following the concept of query-focused summarization [50], which aims to generate summaries based on a particular user’s interest. The query was as follows:

> “Below is the title, journal, and a partial excerpt (introduction + methods) of a biomedical article. Your task is to summarize the article, focusing on the study design.

> Title: *{*title*}*

> Journal: *{*journal title*}* Excerpt: *{*article content*}*”

The article content includes the Introduction and Methods sections, as in other approaches.

### 2.3. PT Classification Models

Our previous best model [32] used a PubMedBERT encoder as the base model. In this work, in an effort to enrich article representations, we evaluated several BERT-based models as base encoders for fine-tuning. Specifically, we experimented with the following BERT-based encoders:

- PubMedBERT [33]: pre-trained from scratch on PubMed abstracts and full-text using masked language modeling and next sentence prediction tasks. It was used in previous work and serves as the baseline.
- BioLinkBERT [34]: pre-trained using a document relation prediction task in lieu of next sentence prediction. It leverages citation links between articles for this task.
- SPECTER [35]: incorporates citation information through a custom contrastive loss that pulls together pairs of article representations if one cites the other; otherwise, representations are pushed apart.
- SPECTER2-Base [36]: extends SPECTER, by pre-training on a larger dataset (x10 as big; 6.2M training triplets across 23 fields of study).
- SPECTER2-Clf [36]: SPECTER2-Base pre-fine-tuned for classification. The model is pre-fine-tuned to predict an article’s field of study (multi-label task) as well as to predict descriptors (e.g., “Brain”,“Breast Neoplasms”) and qualifiers (e.g., “Complications”, “Surgery”) from the 30 most frequent top-level MeSH descriptors for articles with exactly one qualifier (multi-class task). This pre-fine-tuning drives acts similar to contrastive learning, driving article representations from similar fields and descriptors closer/further to improve differentiation between classes.

In our previous work [32], the model was trained using binary cross-entropy loss and AdamW optimizer. This model serves as the baseline in this work. We further experiment with other optimization and regularization techniques to improve the training process and mitigate the effect of potentially noisy data. Specifically, we used RAdam (rectified Adam) [51], which uses warm-up in its implementation and was shown to be less sensitive to hyperparameter changes, making tuning less important. Instead of binary cross-entropy loss, we used asymmetric loss [52], an extension of focal loss [53], which aims to down-weight possibly mislabeled data in addition to down-weighting and hard-thresholding easy negative samples to focus the learning process on hard-to-classify examples. This can lead to faster convergence during training and improved performance when trained on noisy data. Additionally, we used label smoothing [54, 55] to further enhance model robustness against noise as well as to better calibrate the model. By taking a small probability from the correct labels and dispersing it among incorrect labels, label smoothing prevents the model from overfitting its predictions to potentially incorrect labels. This regularization prevents model over-confidence and enhances performance on noisy datasets. We use the default parameters for asymmetric loss: *γ_−_* = 4, *γ*_+_ = 1, *m* = 0.05, and *ϵ* = 1e-8, where *γ* is a weighting term for positive and negative losses, *m* is the probability shifting hyperparameter and *ϵ* is a smoothing term. For label smoothing, we set the *a* parameter, which is the amount of re-distributed probability, to 0.05.

### 2.4. Contrastive Learning

The basic idea behind contrastive learning (CL) is that similar instances (e.g., instances with the same labels) should have representations that are closer within the model embedding space, while dissimilar instances should be further apart. Our previous experiments with CL were inconclusive [32]. In this work, we provide evaluate CL more extensively, experimenting with both unsupervised and supervised CL. We briefly introduce the CL techniques we use below and provide more details about them in Appendix A:

#### Unsupervised CL

We use unsupervised *SimCSE* method [37] as the CL baseline. In this method, an instance’s positive anchor is a dropout-augmented version of itself. Negative anchors are all other instances within the batch.

As negative representations are weighed equally to calculate loss in SimCSE, outliers (or instances that are significantly different) have a greater effect, which may not be optimal. To mitigate this, *ADjusted InfoNCE (AD-NCE)* [38] weighs negative instances using hyperparameters to focus on potentially more informative instances.

#### Supervised CL

While unsupervised CL focuses on improving uniformity (i.e., how evenly distributed all instances are within the representation space), supervised CL focuses on alignment, in which instances with the same label are brought closer together. In multi-label settings, CL is more difficult to implement as similarity is less clear due to the various levels of overlapping labels. To account for this, various contrastive loss functions have been proposed [56, 57], although these sometimes require specialized minibatching processes.

*HeroCon* [39] is a unified CL framework that introduces weighted supervised CL without the need for any specific batching procedures. The loss is weighed through the Hamming distance of each instance’s set of binary labels as a form of supervised similarity. Additionally, HeroCon demonstrates additive benefits when combining both unsupervised and supervised approaches compared to either approach individually.

In supervised CL, HeroCon weights all label differences equally, which may not be optimal in our work. For example, intuitively, representations of cohort studies should probably be more closely related to prospective studies, whereas those of autobiographies should be more related to biographies. Lan et al. [40] introduce *WeighCon*, a supervised contrastive loss which learns optimal weights between labels rather than relying on Hamming distance.

In efforts to improve these supervised contrastive loss functions, we also applied the asymmetric focusing modulation term from asymmetric loss [52] to down-weight easy instances and focus on harder ones, which may be more heterogeneous. We refer to this combination as *noise-aware* supervised CL. Additionally, we experiment with in-batch label correction wherein we consider any instance with a label probability ≥ 0.85 as positively labeled for that class, regardless of its true label, in efforts to better account for false negative labels.

In all experiments with unsupervised and supervised CL, contrastive loss terms were combined with the label loss (i.e., binary cross entropy) by multiplying the CL loss terms with a scaling factor before averaging these losses all together, as shown below:

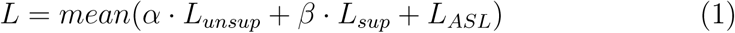

where *α* and *β* are hyperparameters that weigh unsupervised and supervised contrastive loss respectively.

### 2.5. Experimental Setup

We used the following hyperparameters for model training: batch size (32), learning rate (transformer layers): 1e-4, learning rate (classification layer) (1e-2), epochs (25), optimizer (RAdam), dropout (0.1), label smoothing with *α* = 0.05, asymmetric loss with *γ_−_*=4, *γ*_+_=1, and *m*=0.05, and early stopping (when no improvement is observed on validation set macro-F1 over 4 epochs).

Unless the component is being studied or otherwise noted, all experiments used the SPECTER2-Base encoder with the default hyperparameters and *without* label splitting or contrastive loss (CL). This configuration was the strongest non-CL baseline. We did not include the best-performing CL variant (ADNCE + noise-aware HeroCon) in these experiments due to its substantially higher computational cost (approximately 35 minutes per epoch with CL vs. 16 minutes without CL). Experimental settings for CL experiments are provided in Appendix A. All models were implemented using PyTorch (v.2.2.0). Model weights downloaded from the HuggingFace transformers library (v.4.40.2).

All experiments were conducted on a single Tesla v100 GPU with 32GB of memory. Each BERT-based experiment on the larger PubMed dataset took 11.25 hours, while the PMC-based experiments took 1.3 hours each for training. Summary generation for full-text articles required 39.5 hours using PRIMERA, 25 hours using LLaMa, 8.7 hours using BMRetriever, and 7.2 hours using TextRank.

### 2.6. Evaluation

We used standard classification evaluation metrics: precision, recall, and F1 score, as well as area under ROC curve (AUC). Micro- and macro-averaged performance is reported for each metric. Micro-averaged metrics weigh all instances equally, while macro-averaged metrics weigh each class equally. Early stopping was based on macro-averaged F1, and model selection was implemented to maximize macro-averaged F1. This was done instead of loss or micro-F1 to avoid very poor performance on some rare classes. We calculated optimal probabilistic thresholds for labels that had at least 50 positive instances within the validation set; for all others, the threshold was fixed at 0.5. These thresholds were then used to evaluate performance on the test set. Bootstrap sampling on model predictions with 1,000 replicates was used to generate confidence intervals for each experiment (95% CI). Additionally, we assessed model calibration using expected calibration error (ECE) [58], which bins predicted probabilities into k bins (k=15 in this work) and calculates the weighted average of each bin’s difference between the probabilities and the accuracy. We used L2 variant of ECE, which measures the squared difference between predicted probabilities and observed accuracies across confidence bins. We conducted ablation studies to isolate the impact of features and architectural choices (e.g., different base encoders) on model performance. Primary comparisons were conducted using paired significance testing over 1,000 bootstrap replicates of the test set. For each bootstrap sample, models were evaluated on the same resampled instances, and the resulting paired micro-F1 scores were compared using a one-sided paired t-test from the SciPy package (*ttest rel*).

We also calculated core-averaged performance, where “core” consisted of a set of PTs that the authors of this study deemed most important for evidence synthesis. Those PTs were as follows: Case Reports, Case-Control Studies, Clinical Studies as Topic, Clinical Study, Clinical Trial, Clinical Trial Protocol, Cohort Studies, Cross-Over Studies, Cross-Sectional Studies, Double-Blind Method, Evaluation Study, Follow-Up Studies, Longitudinal Studies, Meta-Analysis, Multicenter Study, Prospective Study, Random Allocation, Human Randomized Controlled Trials, Retrospective Studies, Systematic Review, Systematic Reviews as Topic, and Validation Study.

To better understand how the input influences model predictions and enhance model interpretation, we implemented gradient-based saliency mapping [59]. Specifically, we employed Integrated Gradients [60], which assigns importance scores to input features by approximating the integral of the gradients of model outputs with respect to the input. We visualized these scores using the Captum package [61], highlighting keywords that influenced the classification outcome. We compared the model’s predictions with the true labels to gain insights into how these terms contributed to both correct and incorrect predictions.

## 3. Results

### 3.1. Dataset Statistics

Our dataset included 166,192 articles, which were split into train (70%, n = 116,368), validation (10%, n = 16,619), and test (20%, n = 33,205) sets, while preserving the dataset’s overall PT distribution across splits. The distribution of the 61 labels across the dataset is shown in Figure 2 (left panel). Of the 166,192 articles in our dataset, 24,366 were available in the PMC Open Access Subset and were used for full-text experiments: 17,112 in training, 2,420 in validation, and 4,834 in testing.

**Figure 2:**
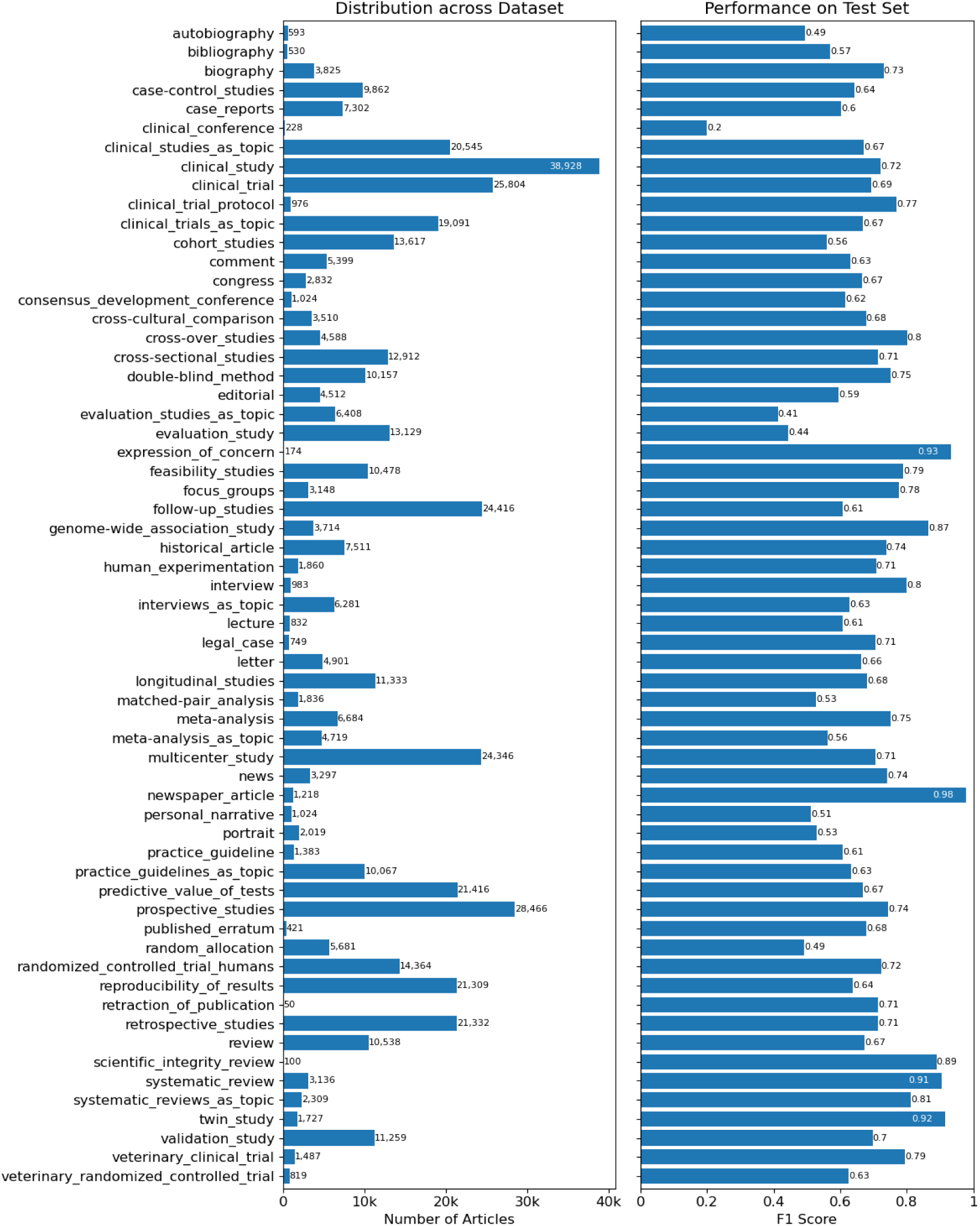
The left panel shows the PT label distribution for all articles in our full dataset. The right panel shows the individual label performances (F1 score) of the best-performing model (SPECTER2-Base with ADNCE and noise-aware HeroCon and using asymmetric loss with label smoothing) on the PubMed test set (n = 33,205).

### 3.2. PT Classification Models with PubMed-only Features

In our primary analysis, we compare the best model and features from our previous work [32] against the best-performing model from this work, which used SPECTER2-Base embeddings, asymmetric loss with label smoothing, and both unsupervised (ADNCE) and supervised contrastive loss (noise-aware HeroCon). The results for these two models are provided in Table 3, which shows an improvement across all metrics except for micro-recall. The results for the best model in this work are also shown in Figure 2 (right panel). This model was significantly better than the model from previous work (both macro-F1 and micro-F1; p < 0.001). Macro-F1 performance on core PTs were also higher in this work (0.691) compared to the previous work (0.686).

**Table 3:** Performance comparison of our previous best model architecture and features [32] with the best model and features from this work. 95% CIs were calculated using bootstrap sampling from the test set (n = 33,205).

| Model | Precision ↑ | | Recall ↑ | | F<sub>1</sub> ↑ | |
| --- | --- | --- | --- | --- | --- | --- |
|  | Micro | Macro | Micro | Macro | Micro | Macro |
| Previous work [32] | 0.637<br>[0.634-0.641] | 0.665<br>[0.660-0.670] | **0.680**<br>[**0.677-0.683**] | 0.641<br>[0.635-0.647] | 0.658<br>[0.655-0.661] | 0.643<br>[0.638-0.648] |
| **This work** | **0.665**<br>[**0.661-0.668**] | **0.684**<br>[**0.675-0.693**] | 0.676<br>[0.673-0.680] | **0.684**<br>[**0.677-0.691**] | **0.670**<br>[**0.668-0.673**] | **0.677**<br>[**0.670-0.684**] |

Ablation study results are reported in Table B.1 (Appendix B). Asymmetric loss variants generally outperformed binary cross entropy variants in terms of F1, while performing worse across all expected calibration error (ECE) metrics. Label smoothing, as expected, lowered ECE, indicating better calibrated models. The citation-informed encoders generally outperformed PubMedBERT. There was no significant difference between the performances of the other models (BioLinkBERT, SPECTER, and SPECTER2 variants), although SPECTER2-Base performed best overall (micro-F1: 0.669; macro-F1: 0.673). Using alternate label splits in training did not improve over the baseline, SPECTER2-Base with no added label splits. In terms of CL methods, SPECTER2-Base model trained with ADNCE and noise-aware HeroCon contrastive losses performed best, significantly better than the baseline. Unsupervised CL approaches (SimCSE, ADNCE) alone underperformed supervised approaches (HeroCon, WeighCon). Incorporating a noise-aware modulation term and in-batch label correction did not improve the performance of supervised CL, while combining noise-aware supervised CL with ADCNE did improve performance somewhat.

### 3.3. PT Classification Models with Full-Text Features

We compared a model trained using PubMed-only features with a model trained using features derived from the full-text in addition to PubMed features. The results of this comparison are shown in Table 4. The best model using full-text features included TextRank, label sentences, first sentence, NCT identifier information, ethics, number features (rough word count of article, # of tables, and # of figures), and primary section heading features. This model outperformed the baseline model using PubMed-only features (title, abstract, and metadata) across both micro- and macro-F1 metrics (p < 0.001). Note that macro-averaged performances of both models are lower than those shown in Table 3, because these experiments used a smaller dataset with full-text articles only (17,112 training instances and 4,834 test instances). Because of this smaller dataset, there were cases where certain PTs had relatively few (or in some cases 0) instances within the validation or test set, so micro-F1 should be given stronger consideration than macro-F1 when comparing full-text models and we do not include macro-F1 results in Table 4.

**Table 4:**
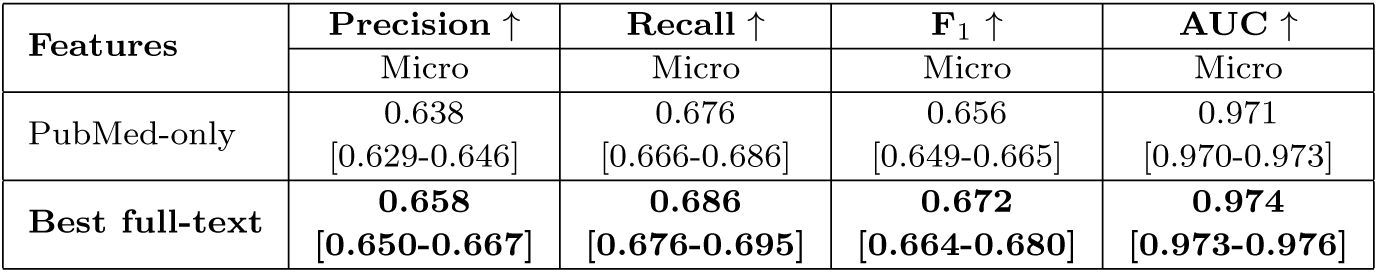
Performance comparison of models trained with PubMed-only features and with the inclusion of best full-text-based features.

We performed ablation studies to better isolate the impact of each feature. The results of these experiments on the full-text test set are provided in Table C.1 in (Appendix C). Removing the first sentence feature (first sentence from the methods section or the article if no methods section present) had the greatest impact on performance. Removing section headings had the least impact on performance. These results are also influenced by the 512-token limit inherent in BERT-based models as this combination of features generally exceeds this limit, requiring truncation.

### 3.4. Impact of Summarization of Full-Text Content

The results for summarization-specific experiments are detailed in Table C.1 (Appendix C), where the PubMed-only features serve as the baseline approach (b). Incorporating summaries across all methods did not enhance micro-F1 compared to the baseline. However, 4 of 6 summaries improved recall, going as high as 0.726 (b + PRIMERA). These results suggest that while summaries do not consistently boost overall performance, they can meaningfully improve the model’s ability to retrieve positive instances, which may be worthwhile in evidence synthesis pipelines.

## 4. Discussion

In this study, we aimed to improve PT classification by improving article representations, including full-text features, creating more homogeneous labels, and accounting for label noise through advanced optimization techniques. Our best model improved on previous work [32], increasing macro-F1 from 0.643 to 0.677 and micro-F1 from 0.658 to 0.670 when evaluated on 61 labels across 33,205 articles. Furthermore, by adding full-text information, we were able to improve model performance on the full-text subset of the test set (n = 4,834) [micro-F1: 0.656 to 0.672; macro-F1 = 0.595 to 0.600]. While performance using the larger PubMed dataset is reasonably high for PTs such as Systematic Review (0.91 F1) and Genome-wide Association Study (0.87), it is lower for some important PTs (e.g., Random Allocation (0.49) and Cohort Studies (0.56)) (Figure 2), indicating that PT classification remains a challenging task.

### 4.1. PT Classification Models with PubMed-only Features

In prior work, we only used PubMed features (title, abstract, and meta-data) as input [32]. Some of our experiments in this study used the same feature set so that we could assess whether enriched article representations, more fine-grained labels, and optimized training could enhance model performance. Our results suggest that a base encoder optimized for citation-aware biomedical document representation without any pre-fine-tuning (SPECTER2-Base) provides enriched representations beneficial for PT classification. Additionally, our hypothesis that citation links between articles could provide additional signal for PT classification did seem to hold true, with citation-aware models (BioLinkBERT, SPECTER, and SPECTER2 variants) generally outperforming PubMedBERT. The largest improvements appear to be in non-research related PTs, e.g., published erratums (PubMedBERT to SPECTER2-Base: +21.0 F1), however, there are some other research oriented PTs that improve as well (PubMedBERT to SPECTER2-Base: letters (+13.5 F1) and systematic reviews (+4.8 F1)). Our “core” metric, which is a macro-average of clinically important PTs, improved slightly as well (core-F1: PubMedBERT (0.686) vs. SPECTER2-Base (0.691)). Future work could explore other auxiliary adaptive pre-training tasks that might benefit PT classification.

Noting the heterogeneity of some PTs (e.g., Cohort Studies, Clinical Study), we also tried to improve the quality of article representations in fine-tuning by splitting labels into more homogeneous classes as well as introducing topical labels for study populations. This was done to explicitly teach the model relationships between labels as well as correlations between populations and study design. However, benefits from this additional knowledge were negligible, which suggests that the model might already be leveraging label correlations in making predictions.

We observed some benefits from optimizing training through the use of asymmetric loss, label smoothing, and CL. We explored asymmetric loss and label smoothing for regularization and mitigating the effect of noisy data on model performance. The preliminary results showed improvements with these optimizations (micro-F1: BCE (0.654) vs. ASL (0.669)). As another added benefit, label smoothing has been shown to calibrate predictions during training [55]. Our results showed similar results with label smoothing comparisons having lower ECE metrics than non-label smoothing experiments. ASL variants had much larger ECE, indicating worse calibration, visualized in Figure B.1 (Appendix B). This is due to the probability shifting in ASL. Future work could aim to address this in order to improve calibration of models using ASL.

Training with CL provided modest performance gains. Models trained with supervised CL approaches led to higher performance overall, while using unsupervised CL alone did not enhance performance. Consistent with findings in computer vision [39], combining unsupervised and supervised CL objectives produced some additional gains. Largest performance gains with CL training came from long-tail PTs, such as Bibliography, Personal Narrative, and Clinical Conference, which often lack abstracts. The best-performing CL configuration (ADNCE + HeroCon + Noise-Aware) more than doubled per-epoch training time compared to training without CL. Given this trade-off, the practical utility of CL for this task may be limited, especially when larger training sets are available.

### 4.2. PT Classification Models with Full-Text Features

Overall, we were able to improve model performance by adding full-text information. The difference in model performance with full-text features was statistically significant, and aligned with our general assumption that detailed information related to study design may not always be present in an article’s abstract. Some features were extracted using simple regular expressions, and further refinements to these expressions could lead to further benefits. For example, a more systematic approach to identify study design information (e.g., information extraction of methodological characteristics [62] rather than simply using sentences with PT label mentions) could be more robust. At the class level, performance improved on certain PTs, perhaps highlighting that information may only be present in the full-text. For example, longitudinal studies (0.718 → 0.754), double-blind method (0.766 → 0.807), multicenter study (0.651 → 0.673), and human RCTs (0.670 → 0.678) all improved when using the best full-text features. We can envision a triage policy that could invoke the full-text model when the PubMed-only model assigns low confidence (e.g., 0.4–0.6) to such classes. This targeted strategy may capture most of the performance gains offered by full-text features while minimizing overall latency and compute costs by avoiding full-text processing when it is unlikely to be beneficial.

The lack of performance improvements from incorporating any summary (extractive, abstractive, or LLM-generated) into our full-text experiments indicates that summaries were ineffective during these initial efforts. Across all variants, models using summaries performed no better than the baseline that relied solely on PubMed-derived features (Table C.1 in Appendix C). This result contrasts with our initial expectations, given that study designs are often more clearly described in the methods section than the abstract and should be accessible through extractive or abstractive techniques. However, several factors likely contributed to the lack of improvement. First, the summaries themselves were often lengthy, exceeding 512 tokens in some cases, with average lengths of 251 tokens for b + primera and 263 tokens for b + llm. Therefore, these summaries were often substantially truncated when appended to the PubMed-only features. Second, because summaries were appended as the final input component, they were the first to be discarded under context-length constraints, reducing their effective contribution. Together, these issues suggest that, in our current setting, summaries did not provide additional discriminative information beyond what was already captured by the PubMed-based model. Future work may explore the effect of feature ordering on performance as well as utilizing encoders with longer input lengths.

Additionally, our abstractive summarization approach relied on off-the-shelf models without task-specific adaptation. Fine-tuning an abstractive model on a dataset explicitly annotated with study design information could potentially enhance performance by reducing hallucinations and paraphrasing inaccuracies. However, to our knowledge, no sufficiently large dataset of this kind currently exists. Still, we believe that summarization could be particularly helpful in three scenarios: (1) when study design details are not explicitly stated but can be inferred from broader contextual clues; (2) when PubMed articles lack abstracts but have full-text; and (3) when the context size exceeds token limits. In these cases, summarization methods may help distill relevant details more effectively.

Despite the improved performance with full-text features, it is important to note that full-text content is often not machine-accessible. Our experiments focused on a relatively small subset of articles with full-text content available on PMC Open Access Subset for XML download. Some articles available in PDF format could be processed with PDF-to-text conversion tools, such as Grobid, and the models could be applied to the extracted text. However, a general solution to classify PTs for all biomedical articles using their full-text remains an open research area. We also note that NLM indexers typically have access to full-text during manual curation.

### 4.3. Error Analysis and Model Interpretation

To better understand how the model makes predictions, we generated visual representations from example instances, highlighting keywords that influenced the classification outcomes. Figure 3 shows the gradient-based saliency maps of two instances. Words in green have a positive association with predicting “True”, while red words have a negative association. As an example of what we would expect, the top panel in Figure 3 shows a correctly predicted instance of Double-Blind Method. In the lower panel, we show an instance with the true label of clinical trials as topic, predicted as a Clinical Trial Protocol. We can observe that the article in fact reports a protocol for a pragmatic RCT. This most likely occurred due to ambiguities within NLM indexing. RCT protocols published before 2019 were only tagged as RCTs instead of as Clinical Trial Protocols [63]. Over time, new PTs have been added or their definitions changed. It is likely that our model is more consistent and accurate compared to NLM indexing, although the performance might still be improved by removing RCT protocol articles from the RCT training set where possible in future work.

**Figure 3:**
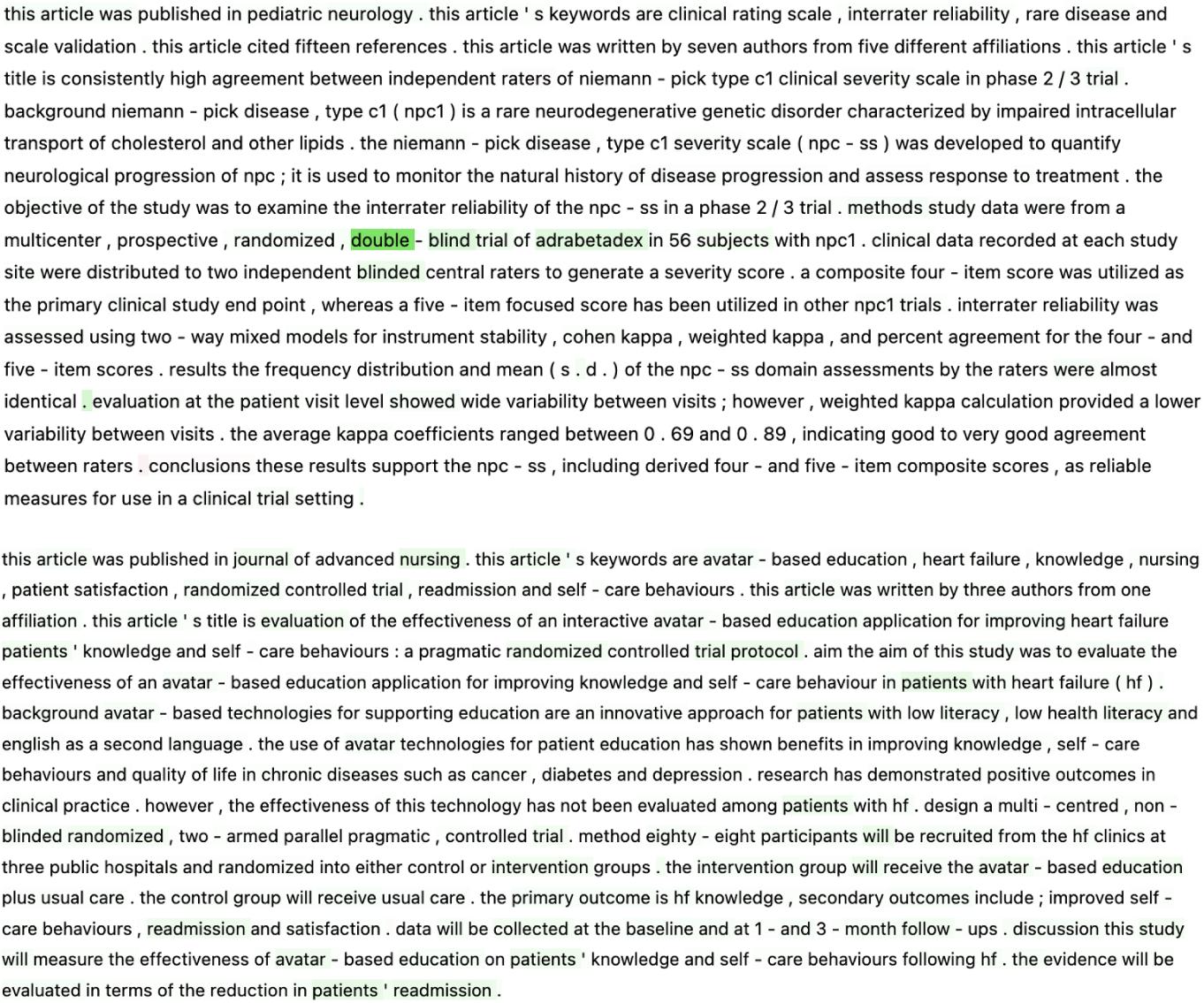
Gradient-based saliency maps for two instances within the test set. Green indicates a positive association with the model predicting a specific label, while red indicates a negative association with a particular label. The first article is PMID: 34952292 and the second is PMID: 29920745.

### 4.4. Dataset Issues

Errors like the one mentioned above raise concerns with the use of PT labels in PubMed for training and evaluation. On one hand, these labels are manually assigned by NLM indexing experts and they are arguably hard to improve upon. These experts are highly trained and follow detailed indexing guidelines with access to the full article text when assigning publication types and MeSH terms. Also given its scale, this dataset is an attractive resource for training broad-coverage PT classification models. On the other hand, previous work highlights the potential noisiness of the PT terms in PubMed [29, 41], which our previous limited manual analyses also confirmed. For this reason, we explicitly treat these labels as silver standard in this work. Conducting a large, fully blinded annotation effort across 61 labels was infeasible within the scope of this work. Instead, to obtain at least a preliminary sense of label quality, we conducted a small exploratory audit of a stratified sample of 333 articles in the test set. One author (HK) conducted the review, which proceeded in two steps: (1) assessing the silver-standard labels, and (2) examining disagreements between best model predictions and silver labels to identify potential labeling noise. Titles, abstracts, and (when available) full-text were used for judgment. The reviewer agreed with the original label sets for 223 of 333 articles (67%) and revised labels for the rest (for a net addition of 36 labels). On this set, the model’s micro-F1 score increased significantly from 0.676 to 0.759 when evaluated against the audited labels. Because this audit was unblinded and per-label sample sizes were too small for reliable error-rate estimation, we do not report noise-adjusted metrics for the full dataset. We do not present this analysis as a substitute for a true blinded evaluation. However, it provides evidence that the dataset contains non-negligible noise and that some model predictions not included in the silver standard are plausible, suggesting that our reported performance likely represents a lower bound on true model performance.

Additional concerns may arise that keyword-based tagging for four PTs (Systematic Review, Systematic Reviews as Topic, Veterinary Clinical Trial, and Veterinary RCT) could lead to pattern-based data leakage. We note that for veterinary PTs, we use keywords only to *exclude* human research, making pattern-based leakage unlikely. To assess leakage for systematic review PTs, we conducted a targeted evaluation on post-2019 articles manually labeled by NLM curators with these PTs. Performance remained strong on these manually annotated subsets (Systematic Review: P=0.966, R=0.974, F1=0.970; Systematic Reviews as Topic: P=0.889, R=0.750, F1=0.814), indicating that keyword filtering did not inflate performance.

Although several smaller, manually annotated datasets exist for PT classification [31, 64], they are limited in size and focus. We took a pragmatic approach and tried to partially address this issue purely as an optimization problem. However, more comprehensive qualitative and quantitative evaluations, ideally involving blinded expert review, are an important direction for future work, both for characterizing labeling noise and for developing methods to improve PT label quality at scale.

### 4.5. Assessment of Model Robustness Under Temporal Distribution Shifts

To partially assess the generalizability of our models to temporal out-of-distribution (OOD) shifts [65, 66, 67, 68], we applied the best-performing model to 198,140 human-labeled articles indexed in 2024, retrieved using the same search strategy as our original dataset but restricted to publication year 2024. We found that performance degraded on this temporally shifted dataset (micro-F1: 0.451; macro-F1: 0.221; AUC: 0.931), in line with other neural modeling works in biomedical NLP [65, 67].

A key factor for this degradation appears to be a substantial shift in label distribution. Compared with the original 1987-2023 dataset, the 2024 sample contains a markedly higher proportion of case reports (1987-2023: 4.4% vs. 2024: 19.8%) and reviews (6.3% vs. 30.4%) and a much lower proportion of clinical studies (23.4% vs. 0.2%). Since the model was trained on a distribution that differs from the 2024 data, this distributional shift likely contributed to the drop in performance. Furthermore, 27 labels had fewer than 30 instances and 3 labels had none, which may explain the especially low macro-F1. The cause of these shifts in label distribution is not entirely clear. One possibility may be that changes in PubMed metadata or indexing workflows in 2024 contributed to the observed differences. For example, although the articles are labeled as neither automatically indexed nor explicitly human-curated, it is plausible that many have not yet undergone the full human indexing process. However, we cannot confirm this with the available metadata. A more systematic investigation of temporal, topical, journal-level, and other domain shifts in publication type and study design tagging represents an important direction for future research.

### 4.6. Comparison with In-Context Learning

While previous evaluations of autoregressive LLMs (e.g., GPT and LLaMA) in BioNLP tasks have shown that, in zero- and few-shot settings, they are typically outperformed by traditional fine-tuning with BERT models [69], it is still important to evaluate these models empirically as they offer a flexible baseline. To assess how LLMs perform in our setting compared to a fine-tuned BERT model, we implemented a zero-shot baseline using GPT-4o (gpt-4o-2024-08-06) and LLaMA-70B (meta-llama/llama-3.3-70B-instruct) as representatives of proprietary and open-weight models, respectively. Both models were prompted with a manually constructed zero-shot template (see Appendix D for the full prompt).

For a fair comparison, we provided the LLMs with the same PubMed-only features used by the BERT-based model, including the article’s title, abstract, and structured metadata (e.g., keywords, chemical names, and journal), and instructed them to return all applicable labels based on the predefined label definitions. GPT-4o achieved a macro-F1 of 0.430 and a micro-F1 of 0.476, while LLaMA-70B achieved a macro-F1 of 0.328 and a micro-F1 of 0.364; both falling short of the BERT-based model evaluated on the full-text subset of the test set (macro-F1: 0.592; micro-F1: 0.656). Additionally, inference runtime for LLM experiments (LLaMa-70B: 4.73 hours; GPT-4o: 2.83 hours) were significantly longer than that of the BERT-based model (45 seconds), which may limit the feasibility of LLMs as indexers given the high publication rate of biomedical literature.

Analysis revealed that LLMs overpredicted labels, particularly for articles with no annotated publication types (30% of the test set), leading to false positives and reduced precision (GPT-4o: 0.476; LLaMA-70B: 0.288). This behavior is consistent with a known limitation of LLMs: their tendency toward overconfidence in zero-shot classification settings without task-specific calibration or fine-tuning [70]. In addition, the models struggled with fine-grained distinctions, such as differentiating Clinical Study from Clinical Trial, or distinguishing “as topic” labels from actual study reports.

### 4.7. Limitations

There are several limitations related to this study. The main challenge relates to evaluation due to non-negligible noise in the dataset, as discussed above. Despite acknowledging the issue and trying to mitigate it using optimization techniques in training, we still need to use a somewhat noisy test set for our evaluation. However, our exploratory audit suggests that model performance could be underestimated due to the noise. Future work needs to better understand the patterns of noise in the PubMed PTs and try to establish a true gold standard, at least for evaluation, although this seems very challenging at large scale. Second, while our list of publication types and study designs is extensive, it is not exhaustive. Our current model does not classify some PTs initially deemed to have relatively little utility within the biomedical community (e.g., Directory, Journal Article). At the same time, PubMed PTs are also not exhaustive (e.g., Case Series is not a PubMed PT). In future work, we aim to expand our classification scheme to include all PTs indexed in PubMed as well as new PTs that would serve unmet needs of the diverse biomedical research community.

Our initial explorations with zero-shot learning using autoregressive LLMs yielded relatively poor results. Future work could explore few-shot prompting, instruction tuning, or lightweight adaptation strategies to bridge this performance gap.

## 5. Conclusions

In this study, we trained and validated Transformer-based models for PT classification on a dataset of biomedical articles labeled with 61 PTs using a combination of PubMed queries and indexing terms. Specifically, we compared different base encoders for article representation, investigated whether more fine-grained labels created through label splitting and injecting relevant MeSH terms could enhance performance, and experimented with regularization and optimization techniques for enhanced training, reducing overfitting, and mitigating label noise to some extent. Additionally, we investigated the use of full-text features within PT classification. Our model performance improves upon that reported in previous work [32]; in particular, our results demonstrate the value of incorporating enriched article representations and full-text information into automated PT classification models. This model, while imperfect, could help to ensure consistent PT indexing within repositories like PubMed and beyond when deployed (e.g., Semantic Scholar, etc.). Future work will analyze model output more extensively through manual analysis, continue to expand to include PTs not currently considered, as well as deploy these models into existing systems (e.g., Anne O’Tate [71]) to increase accessibility to both academia and research repositories. We will also explore autoregressive LLMs with larger context sizes in more depth for PT classification.

## Data Availability

Data, code, and models are available at https://github.com/ScienceNLP-Lab/MultiTagger-v2.

## Acknowledgments

This work was supported by the National Library of Medicine of the National Institutes of Health under the award number R01LM14292. The content is solely the responsibility of the authors and does not necessarily represent the official views of the National Institutes of Health. The funder had no role in considering the study design or in the collection, analysis, interpretation of data, writing of the report, or decision to submit the article for publication.

## Author Contributions

Joe D. Menke: Conceptualization, Methodology, Software, Visualization, Validation, Formal Analysis, Data Curation, Writing – original draft, and Writing – review and editing; Shufan Ming: Conceptualization, Methodology, Software, Visualization, Writing – original draft, and Writing – review and editing; Shruthan Radhakrishna: Software; Halil Kilicoglu: Conceptualization, Funding acquisition, Methodology, Data Curation, Supervision, Writing – original draft, and Writing – review and editing; Neil R. Smalheiser: Conceptualization, Funding acquisition, Supervision, and Writing – review and editing.

## Appendix A. Contrastive Learning Methods

*SimCSE [37].* Unsupervised contrastive loss in SimCSE is calculated as follows:

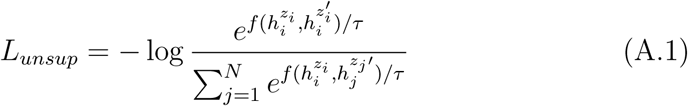

where *N* is the number of instances within the batch, *h^zi^* is the hidden representation of the [CLS] token of an instance, *i*, *h^zi′^* is a dropout-augmented representation, *f* (.,.) is a similarity measurement function (in this case, cosine similarity), and *τ* is a temperature hyperparameter.

*ADjusted InfoNCE (ADNCE) [38].* ADNCE weighs negative instances using hyperparameters to focus on potentially more informative instances. This is done by Gaussian-like weighting negative anchors:

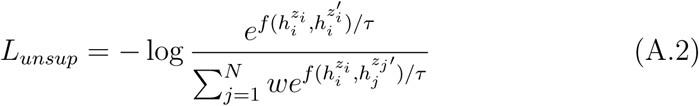

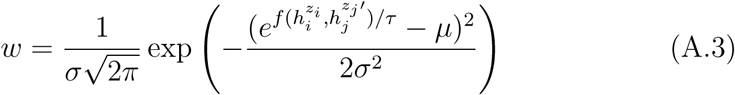

where *σ* controls the weight discrepancy among the samples and *µ* controls the region of weight allocation (i.e., samples closer to *µ* have larger weights).

*HeroCon [39].* In HeroCon,the contrastive loss is weighed through the Ham-ming distance of each instance’s set of binary labels as a form of supervised similarity. This is shown below:

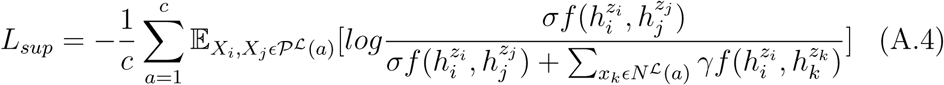

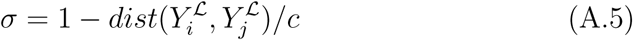

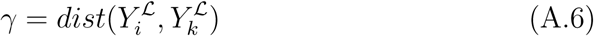

where *dist*(*Y ^L^, Y ^L^*) is used to weigh label similarity (i.e., Hamming distance), *^L^*(*a*) = *X_j_ Y ^L^*(*a*) = 1 is the set of positive instances with the *a^th^* label, and *^L^*(*a*) = *X_k_ Y ^L^*(*a*) = 1 is the set of negative instances. Here, *a* is a PT (e.g., articles in a batch positively labeled as RCTs), *c* is the number of labels, and *γ* is the label similarity weighting, in this case, Hamming distance.

*WeighCon [40].* WeighCon is a supervised CL method which learns optimal weights between labels rather than relying on Hamming distance. This follows the same formulation as above except with a learned weight:

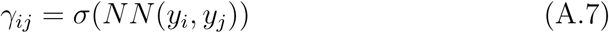

where *σ* is a sigmoid activation function and *NN* is a one-layer, fully connected linear layer, taking in two label vectors and outputting a scalar value corresponding to the similarity between label vectors.

### Noise-aware weighting

In efforts to improve these supervised CL functions, we also applied the asymmetric focusing modulation term from asymmetric loss [52], as shown below:

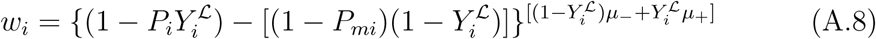

where *P_i_* is the sigmoid activated model prediction for a particular label in instance *i*, *Y ^L^* is the true label, *P_m_i* is the probability after asymmetric clipping (*P_mi_* = *max*(*P_i_* + *m,* 1)), and *µ_−_* and *µ*_+_ are focusing parameters. We use default values *m*=0.05, *µ_−_*=4 and *µ*_+_=1. Overall, the weighting term *w_i_* is used to modulate the HeroCon and WeighCon weighting terms defined in equations A.5, A.6, and A.7 to downweight easy instances and focus on harder ones, which may be more heterogeneous.

### Experimental Settings for Contrastive Learning

All CL experiments used the same temperature parameter (*τ* = 0.05). We also set both ADNCE hyperparameters *µ* and *σ* to 1.0. For HeroCon and WeighCon experiments, we used the hyperparameters that Zheng et al. [39] used with the CelebA dataset, a multi-label dataset with 40 labels that is most similar to ours evaluated in that work. Unsupervised CL experiments used *α* = 0.01, supervised CL experiments used *β* = 0.1, and combined approaches used *α* = 0.1 and *β* = 0.1.

## Appendix B. Ablation studies and analyses with PubMed-only features

Table B.1 shows the results of all PubMed ablation studies.

**Table B.1:**
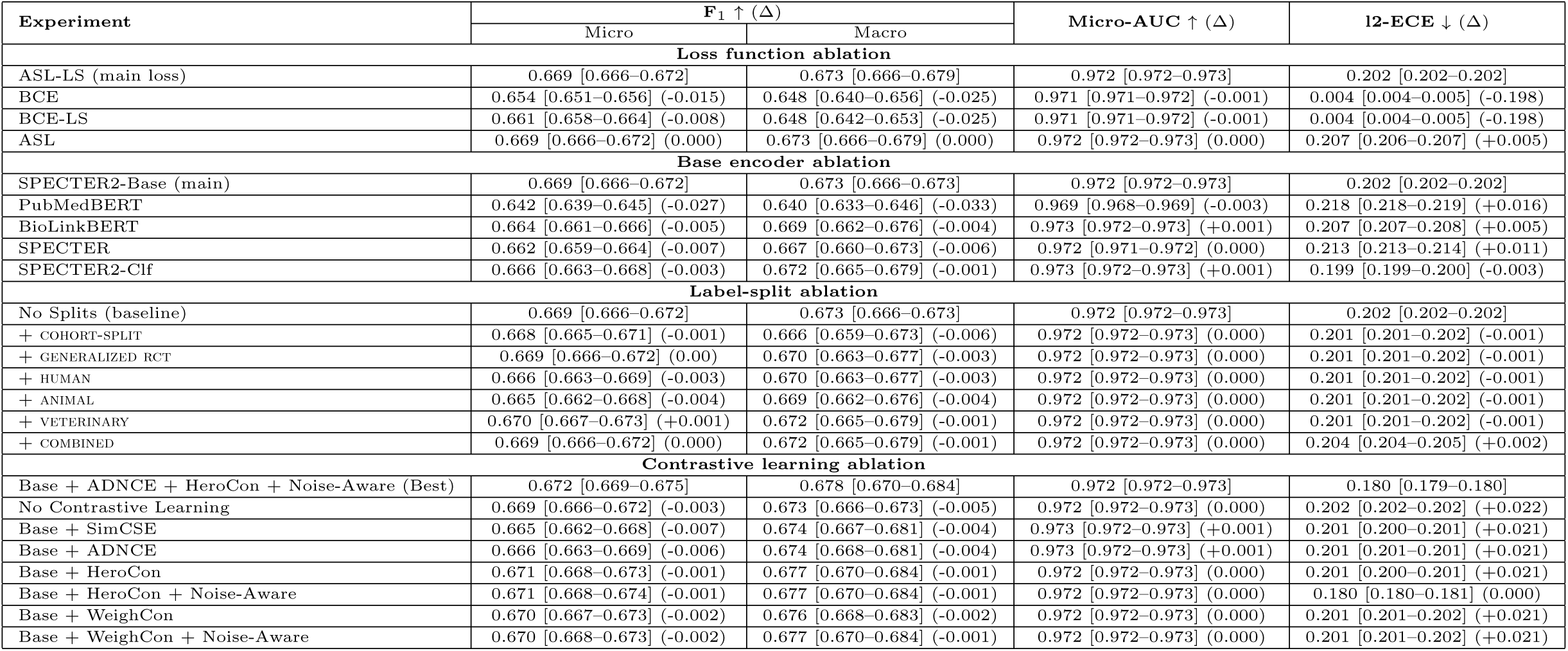
Ablation results for PubMed-only models. Δ shows the performance differences from the main models.

### Appendix B.1. Loss functions and calibration

Figure B.1 plots the probability distribution of model predictions for each loss function. In a well-calibrated model, 90% of instances predicted with 0.9 would be positively labeled; 10% of instances with 0.1 would be positively labeled. With ASL, focusing results in a model that is more poorly calibrated than BCE. Despite this, the overall performance, including AUC (micro-AUC [95% CI]: BCE=0.973 [0.973-0.974], ASL=0.974 [0.974-0.975]) is better when using ASL. To ensure a high recall (sensitivity) while limiting false positives, optimal thresholds should be determined empirically rather than simply using low probability cut-offs (0.01) as is more common in well-calibrated models.

**Figure B.**
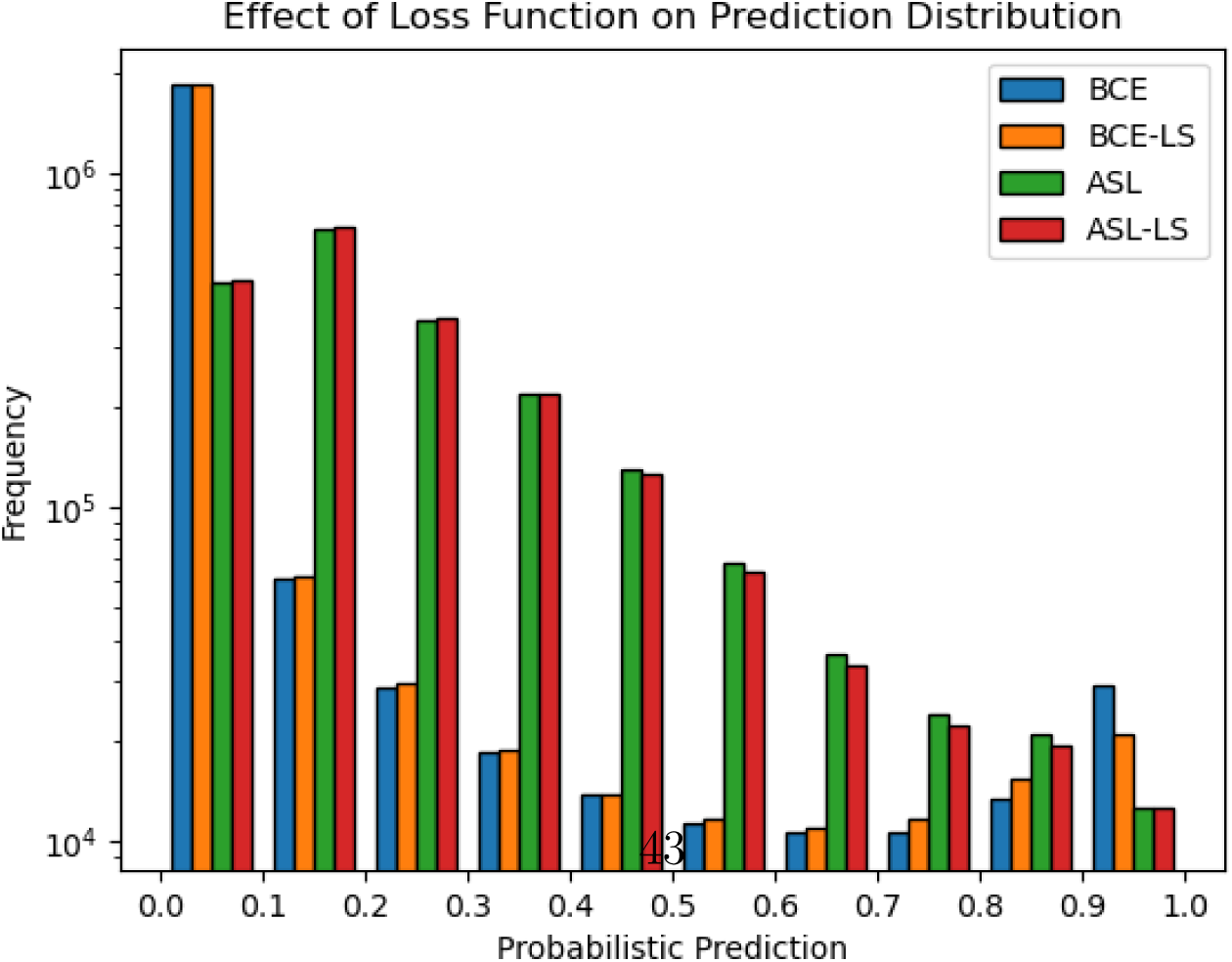
1: The distribution of probabilistic predictions for articles in the test set for models in the loss function ablation study, which evaluates binary cross entropy, binary cross entropy with label smoothing, asymmetric loss, and asymmetric loss with label smoothing.

## Appendix C. Ablation studies and analyses with full-text features

Figure C.1 shows the PT label distribution for the full-text dataset and the performances of the base model and the best-performing full-text model on the full-text test set.

**Figure C.1:**
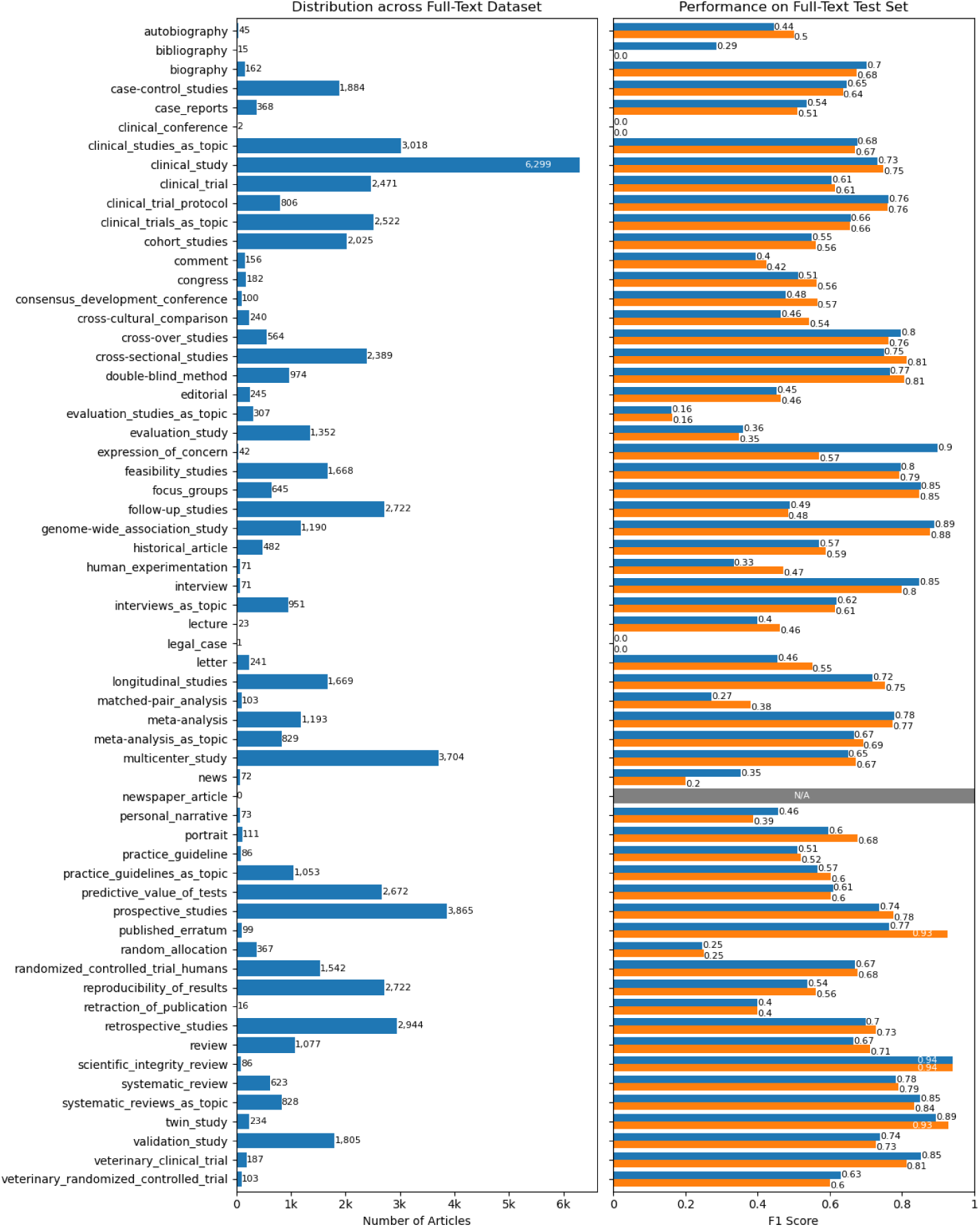
The left panel shows the PT label distribution for articles in our full-text dataset. The right panel shows the individual label performances (F1 score) of the base model (blue) and the best-performing full-text model (orange) on the full-text test set (n = 4,834). The base model uses no full-text features, just features derived from PubMed (e.g., title, abstract, etc.). The best model uses the following full-text features: extractive (multiple), label sentences, first sentence, NCT identifier information, ethics, number features (rough word count of article, # of tables, and # of figures), and primary section heading features.

Table C.1 shows the results of the ablation study of different combinations of full-text features and the performance comparison of models using full-text features and summarization techniques.

**Table C.1:** Ablation scoreboard summarizing changes (Δ) in full-text performance relative to the best combination of features for the ablation experiments and the PubMed features only baseline (B) for the summary experiments. Each cell is intended to show the difference in metric value, a 95% confidence interval, and a delta indicating the absolute difference from the comparator.

| Experiment | F<sub>1</sub> ↑ (Δ) |  | Micro-AUC ↑ (Δ) | ECE ↓ (Δ) |
| --- | --- | --- | --- | --- |
|  | Micro | Macro |  |  |
| Full-text feature ablation |  |  |  |  |
| Best Combination | 0.672 [0.664–0.680] | 0.600 [0.576–0.625] | 0.974 [0.973–0.974] | 0.187 [0.185–0.188] |
| - First sentence | 0.660 [0.653–0.668] (-0.012) | 0.596 [0.571–0.619] (-0.004) | 0.974 [0.973–0.975] (0.000) | 0.187 [0.185–0.189] (0.000) |
| - Label sentences | 0.667 [0.660–0.675] (-0.005) | 0.599 [0.576–0.623] (-0.001) | 0.974 [0.972–0.975] (0.000) | 0.189 [0.187–0.191] (+0.002) |
| - Ethics | 0.664 [0.656–0.671] (-0.008) | 0.597 [0.573–0.620] (-0.003) | 0.974 [0.973–0.975] (0.000) | 0.193 [0.192–0.195] (+0.006) |
| - NCT identifiers | 0.668 [0.661–0.676] (-0.004) | 0.597 [0.573–0.620] (-0.003) | 0.974 [0.973–0.976] (0.000) | 0.187 [0.186–0.189] (0.000) |
| - Number features | 0.669 [0.662–0.677] (-0.003) | 0.600 [0.577–0.614] (0.000) | 0.974 [0.973–0.976] (0.000) | 0.187 [0.185–0.189] (0.000) |
| - Section headings | 0.670 [0.662–0.678] (-0.002) | 0.594 [0.570–0.617] (-0.006) | 0.974 [0.973–0.975] (0.000) | 0.187 [0.185–0.189] (0.000) |
| - TextRank | 0.662 [0.654–0.670] (-0.010) | 0.594 [0.572–0.616] (-0.006) | 0.974 [0.973–0.975] (0.000) | 0.186 [0.184–0.188] (-0.001) |
| Summarization and full-text feature models (vs. B) |  |  |  |  |
| PubMed features only (B) | 0.656 [0.649–0.665] | 0.595 [0.571–0.618] | 0.971 [0.970–0.973] | 0.189 [0.188–0.191] |
| B + TextRank | 0.652 [0.645–0.660] (-0.004) | 0.586 [0.563–0.611] (-0.009) | 0.972 [0.970–0.973] (+0.001) | 0.190 [0.189–0.192] (+0.001) |
| B + EXTRACTIVE (SINGLE) | 0.654 [0.646–0.662] (-0.002) | 0.584 [0.559–0.609] (-0.011) | 0.972 [0.970–0.973] (+0.001) | 0.190 [0.188–0.191] (+0.001) |
| B + EXTRACTIVE (MULTIPLE) | 0.651 [0.634–0.659] (-0.005) | 0.584 [0.559–0.607] (-0.011) | 0.972 [0.970–0.974] (+0.001) | 0.195 [0.193–0.197] (+0.006) |
| B + EXTRACTIVE (MULTIPLE+DEFINITION) | 0.651 [0.643–0.659] (+0.005) | 0.585 [0.562–0.607] (+0.010) | 0.972 [0.971–0.974] (+0.001) | 0.190 [0.188–0.192] (+0.001) |
| B + LLM | 0.655 [0.647–0.663] (-0.001) | 0.577 [0.555–0.599] (-0.018) | 0.973 [0.972–0.975] (+0.002) | 0.191 [0.189–0.192] (+0.002) |
| B + PRIMERA | 0.654 [0.646–0.661] (-0.002) | 0.570 [0.546–0.594] (-0.025) | 0.972 [0.971–0.974] (+0.001) | 0.205 [0.204–0.207] (+0.016) |

## Appendix D. Prompt Template for LLMs

### System Prompt

You are a biomedical research assistant tasked with identifying the publication type(s) and study design(s) of a biomedical journal article. This is a multi-label classification task. You must select ALL applicable labels that describe the study’s purpose, structure, and design.

Instructions:

- Only use labels from the list provided.
- Do NOT invent new labels. If multiple labels apply, include all of them.
- If no labels apply, output an empty list.
- Your final answer must be in Python list format, as shown below. Label definitions:

### List of label definitions

Answer Format (required):

Labels: [label1, label2, …]

Example:

Labels: [clinical_study, cohort_studies, follow-up_studies]

### Human Input

Title, abstract, and structured metadata of the article (i.e., PubMed-only features)

## References

[1] D. L. Sackett, W. M. Rosenberg, J. M. Gray, R. B. Haynes, W. S. Richardson, Evidence based medicine: what it is and what it isn’t, BMJ 312 (7023) (1996) 71–72.

[2] A. M. Cohen, C. E. Adams, J. M. Davis, C. Yu, P. S. Yu, W. Meng, L. Duggan, M. McDonagh, N. R. Smalheiser, Evidence-based medicine, the essential role of systematic reviews, and the need for automated text mining tools, in: Proceedings of the 1st ACM International Health Informatics Symposium, 2010, pp. 376–380.

[3] S. Khangura, K. Konnyu, R. Cushman, J. Grimshaw, D. Moher, Evidence summaries: the evolution of a rapid review approach, Systematic Reviews 1 (1) (2012) 1–9.

[4] J. Clark, P. Glasziou, C. Del Mar, A. Bannach-Brown, P. Stehlik, A. M. Scott, A full systematic review was completed in 2 weeks using automation tools: a case study, Journal of Clinical Epidemiology 121 (2020) 81–90.

5. National Library of Medicine (US), Incorporating Values for Indexing Method in MEDLINE/PubMed XML, NLM Tech Bulletin e2 (2018) 423, accessed on 02.26.2024. URL nlm.nih.gov/pubs/techbull/ja18/ja18\_indexing\_method.html

[6] A. R. Aronson, J. G. Mork, C. W. Gay, S. M. Humphrey, W. J. Rogers, The NLM indexing initiative’s medical text indexer, in: MEDINFO 2004, IOS Press, 2004, pp. 268–272.

7. J. G. Mork, A. Jimeno-Yepes, A. R. Aronson, et al., The NLM Medical Text Indexer System for Indexing Biomedical Literature, BioASQ@ CLEF 1 (2013).

[8] J. Mork, A. Aronson, D. Demner-Fushman, 12 years on–Is the NLM medical text indexer still useful and relevant?, Journal of Biomedical Semantics 8 (1) (2017) 1–10.

[9] A. R. Rae, J. G. Mork, D. Demner-Fushman, A Neural Text Ranking Approach for Automatic MeSH Indexing., in: CLEF (Working Notes), 2021, pp. 302–312.

10. National Library of Medicine (US), Frequently Asked Questions about Indexing for MEDLINE, accessed on 02.26.2024 (2010). URL nlm.nih.gov/bsd/indexfaq.html

[11] G. Tsatsaronis, G. Balikas, P. Malakasiotis, I. Partalas, M. Zschunke, M. R. Alvers, D. Weissenborn, A. Krithara, S. Petridis, D. Polychronopoulos, et al., An overview of the BIOASQ large-scale biomedical semantic indexing and question answering competition, BMC Bioinformatics 16 (1) (2015) 1–28.

[12] A. Nentidis, A. Krithara, K. Bougiatiotis, M. Krallinger, C. Rodriguez-Penagos, M. Villegas, G. Paliouras, Overview of BioASQ 2020: The eighth bioASQ challenge on large-scale biomedical semantic indexing and question answering, in: Experimental IR Meets Multilinguality, Multimodality, and Interaction: 11th International Conference of the CLEF Association, CLEF 2020, Thessaloniki, Greece, September 22–25, 2020, Proceedings 11, Springer, 2020, pp. 194–214.

[13] A. Nentidis, G. Katsimpras, A. Krithara, S. Lima López, E. Farŕe-Maduell, L. Gasco, M. Krallinger, G. Paliouras, Overview of BioASQ 2023: The eleventh bioASQ challenge on large-scale biomedical semantic indexing and question answering, in: International Conference of the Cross-Language Evaluation Forum for European Languages, Springer, 2023, pp. 227–250.

[14] K. Liu, S. Peng, J. Wu, C. Zhai, H. Mamitsuka, S. Zhu, MeSHLabeler: improving the accuracy of large-scale MeSH indexing by integrating diverse evidence, Bioinformatics 31 (12) (2015) i339–i347.

[15] S. Peng, R. You, H. Wang, C. Zhai, H. Mamitsuka, S. Zhu, DeepMeSH: deep semantic representation for improving large-scale MeSH indexing, Bioinformatics 32 (12) (2016) i70–i79.

[16] R. You, Y. Liu, H. Mamitsuka, S. Zhu, BERTMeSH: deep contextual representation learning for large-scale high-performance MeSH indexing with full text, Bioinformatics 37 (5) (2021) 684–692.

[17] S. Dai, R. You, Z. Lu, X. Huang, H. Mamitsuka, S. Zhu, FullMeSH: improving large-scale MeSH indexing with full text, Bioinformatics 36 (5) (2020) 1533–1541.

[18] G. Xun, K. Jha, Y. Yuan, Y. Wang, A. Zhang, MeSHProbeNet: a self-attentive probe net for MeSH indexing, Bioinformatics 35 (19) (2019) 3794–3802.

[19] X. Wang, R. Mercer, F. Rudzicz, KenMeSH: Knowledge-enhanced End-to-end Biomedical Text Labelling, in: S. Muresan, P. Nakov, A. Villavicencio (Eds.), Proceedings of the 60th Annual Meeting of the Association for Computational Linguistics (Volume 1: Long Papers), Association for Computational Linguistics, Dublin, Ireland, 2022, pp. 2941–2951. doi:10.18653/v1/2022.acl-long.210.

[20] K. Knight, S. Wade, L. Balducci, Prevalence and outcomes of anemia in cancer: a systematic review of the literature, The American Journal of Medicine 116 (7) (2004) 11–26.

[21] N. L. Wilczynski, R. B. Haynes, Consistency and accuracy of indexing systematic review articles and meta-analyses in medline, Health Information & Libraries Journal 26 (3) (2009) 203–210.

[22] L. Peirson, D. Fitzpatrick-Lewis, D. Ciliska, R. Warren, Screening for cervical cancer: a systematic review and meta-analysis, Systematic Reviews 2 (2013) 1–14.

[23] X. I. Yao, X. Wang, P. J. Speicher, E. S. Hwang, P. Cheng, D. H. Harpole, M. F. Berry, D. Schrag, H. H. Pang, Reporting and guidelines in propensity score analysis: a systematic review of cancer and cancer surgical studies, JNCI: Journal of the National Cancer Institute 109 (8) (2017) djw323.

[24] A. M. Cohen, N. R. Smalheiser, M. S. McDonagh, C. Yu, C. E. Adams, J. M. Davis, P. S. Yu, Automated confidence ranked classification of randomized controlled trial articles: an aid to evidence-based medicine, Journal of the American Medical Informatics Association 22 (3) (2015) 707–717.

[25] B. C. Wallace, A. Noel-Storr, I. J. Marshall, A. M. Cohen, N. R. Smalheiser, J. Thomas, Identifying reports of randomized controlled trials (RCTs) via a hybrid machine learning and crowdsourcing approach, Journal of the American Medical Informatics Association 24 (6) (2017) 1165–1168.

[26] I. J. Marshall, A. Noel-Storr, J. Kuiper, J. Thomas, B. C. Wallace, Machine learning for identifying randomized controlled trials: an evaluation and practitioner’s guide, Research Synthesis Methods 9 (4) (2018) 602–614.

[27] N. R. Smalheiser, A. M. Cohen, Design of a generic, open platform for machine learning-assisted indexing and clustering of articles in PubMed, a biomedical bibliographic database, Data and Information Management 2 (1) (2018) 27–36.

[28] A. M. Cohen, J. Schneider, Y. Fu, M. S. McDonagh, P. Das, A. W. Holt, N. R. Smalheiser, Fifty ways to tag your pubtypes: Multitagger, a set of probabilistic publication type and study design taggers to support biomedical indexing and evidence-based medicine, medRxiv (2021) 2021–07.

[29] J. Schneider, L. Hoang, Y. Kansara, A. M. Cohen, N. R. Smalheiser, Evaluation of publication type tagging as a strategy to screen randomized controlled trial articles in preparing systematic reviews, JAMIA Open 5 (1) (2022) ooac015.

[30] R. Proescholdt, T.-K. Hsiao, J. Schneider, A. M. Cohen, M. S. McDonagh, N. R. Smalheiser, Testing a filtering strategy for systematic reviews: evaluating work savings and recall, AMIA Summits on Translational Science Proceedings 2022 (2022) 406.

[31] M. Neves, A. Klippert, F. Knöspel, J. Rudeck, A. Stolz, Z. Ban, M. Becker, K. Diederich, B. Grune, P. Kahnau, et al., Automatic classification of experimental models in biomedical literature to support searching for alternative methods to animal experiments, Journal of Biomedical Semantics 14 (1) (2023) 13.

[32] J. Menke, H. Kilicoglu, N. Smalheiser, Publication type tagging using transformer models and multi-label classification, in: AMIA Annual Symposium Proceedings, American Medical Informatics Association, 2024.

[33] Y. Gu, R. Tinn, H. Cheng, M. Lucas, N. Usuyama, X. Liu, T. Naumann, J. Gao, H. Poon, Domain-specific language model pretraining for biomedical natural language processing, ACM Transactions on Computing for Healthcare (HEALTH) 3 (1) (2021) 1–23.

[34] M. Yasunaga, J. Leskovec, P. Liang, LinkBERT: Pretraining Language Models with Document Links, in: S. Muresan, P. Nakov, A. Villavicencio (Eds.), Proceedings of the 60th Annual Meeting of the Association for Computational Linguistics (Volume 1: Long Papers), Association for Computational Linguistics, Dublin, Ireland, 2022, pp. 8003–8016. doi:10.18653/v1/2022.acl-long.551. URL aclanthology.org/2022.acl-long.551

[35] A. Cohan, S. Feldman, I. Beltagy, D. Downey, D. Weld, SPECTER: Document-level Representation Learning using Citation-informed Transformers, in: D. Jurafsky, J. Chai, N. Schluter, J. Tetreault (Eds.), Proceedings of the 58th Annual Meeting of the Association for Computational Linguistics, Association for Computational Linguistics, Online, 2020, pp. 2270–2282. doi:10.18653/v1/2020.acl-main.207. URL aclanthology.org/2020.acl-main.207

[36] A. Singh, M. D’Arcy, A. Cohan, D. Downey, S. Feldman, SciRepEval: A Multi-Format Benchmark for Scientific Document Representations, in: H. Bouamor, J. Pino, K. Bali (Eds.), Proceedings of the 2023 Conference on Empirical Methods in Natural Language Processing, Association for Computational Linguistics, Singapore, 2023, pp. 5548–5566. doi:10.18653/v1/2023.emnlp-main.338. URL aclanthology.org/2023.emnlp-main.338

[37] T. Gao, X. Yao, D. Chen, SimCSE: Simple Contrastive Learning of Sentence Embeddings, in: M.-F. Moens, X. Huang, L. Specia, S. W.-t. Yih (Eds.), Proceedings of the 2021 Conference on Empirical Methods in Natural Language Processing, Association for Computational Linguistics, Online and Punta Cana, Dominican Republic, 2021, pp. 6894–6910.

[38] J. Wu, J. Chen, J. Wu, W. Shi, X. Wang, X. He, Understanding contrastive learning via distributionally robust optimization, Advances in Neural Information Processing Systems 36 (2024).

[39] L. Zheng, J. Xiong, Y. Zhu, J. He, Contrastive learning with complex heterogeneity, in: Proceedings of the 28th ACM SIGKDD Conference on Knowledge Discovery and Data Mining, 2022, pp. 2594–2604.

[40] M. Lan, L. Zheng, S. Ming, H. Kilicoglu, Multi-label Sequential Sentence Classification via Large Language Model, in: Findings of the Association for Computational Linguistics: EMNLP 2024, 2024, pp. 16086–16104.

[41] A. M. Cohen, Z. O. Dunivin, N. R. Smalheiser, A probabilistic automated tagger to identify human-related publications, Database 2018 (2018) bay079.

[42] N. F. Liu, K. Lin, J. Hewitt, A. Paranjape, M. Bevilacqua, F. Petroni, P. Liang, Lost in the middle: How language models use long contexts, Transactions of the Association for Computational Linguistics 12 (2024) 157–173.

[43] J. Maynez, S. Narayan, B. Bohnet, R. McDonald, On Faithfulness and Factuality in Abstractive Summarization, in: D. Jurafsky, J. Chai, N. Schluter, J. Tetreault (Eds.), Proceedings of the 58th Annual Meeting of the Association for Computational Linguistics, Association for Computational Linguistics, Online, 2020, pp. 1906–1919. doi:10.18653/v1/2020.acl-main.173. URL aclanthology.org/2020.acl-main.173/

[44] R. Mihalcea, P. Tarau, TextRank: Bringing Order into Text, in: D. Lin, D. Wu (Eds.), Proceedings of the 2004 Conference on Empirical Methods in Natural Language Processing, Association for Computational Linguistics, Barcelona, Spain, 2004, pp. 404–411. URL aclanthology.org/W04-3252/

[45] R. Xu, W. Shi, Y. Yu, Y. Zhuang, Y. Zhu, M. D. Wang, J. C. Ho, C. Zhang, C. Yang, BMRetriever: Tuning Large Language Models as Better Biomedical Text Retrievers, in: Y. Al-Onaizan, M. Bansal, Y.-N. Chen (Eds.), Proceedings of the 2024 Conference on Empirical Methods in Natural Language Processing, Association for Computational Linguistics, Miami, Florida, USA, 2024, pp. 22234–22254. doi:10.18653/v1/2024.emnlp-main.1241. URL aclanthology.org/2024.emnlp-main.1241/

46. I. Beltagy, M. E. Peters, A. Cohan, Longformer: The long-document transformer, arXiv preprint arXiv:2004.05150 (2020).

[47] A. Cohan, F. Dernoncourt, D. S. Kim, T. Bui, S. Kim, W. Chang, N. Goharian, A Discourse-Aware Attention Model for Abstractive Summarization of Long Documents, in: M. Walker, H. Ji, A. Stent (Eds.), Proceedings of the 2018 Conference of the North American Chapter of the Association for Computational Linguistics: Human Language Technologies, Volume 2 (Short Papers), Association for Computational Linguistics, New Orleans, Louisiana, 2018, pp. 615–621. doi:10.18653/v1/N18-2097. URL aclanthology.org/N18-2097/

[48] W. Xiao, I. Beltagy, G. Carenini, A. Cohan, PRIMERA: Pyramid-based Masked Sentence Pre-training for Multi-document Summarization, in: S. Muresan, P. Nakov, A. Villavicencio (Eds.), Proceedings of the 60th Annual Meeting of the Association for Computational Linguistics (Volume 1: Long Papers), Association for Computational Linguistics, Dublin, Ireland, 2022, pp. 5245–5263. doi:10.18653/v1/2022.acl-long.360. URL aclanthology.org/2022.acl-long.360/

[49] A. Dubey, A. Jauhri, A. Pandey, A. Kadian, A. Al-Dahle, A. Letman, A. Mathur, A. Schelten, A. Yang, A. Fan, et al., The Llama 3 Herd of Models, arXiv preprint arXiv:2407.21783 (2024).

[50] J. Vig, A. Fabbri, W. Kryscinski, C.-S. Wu, W. Liu, Exploring Neural Models for Query-Focused Summarization, in: M. Carpuat, M.-C. de Marneffe, I. V. Meza Ruiz (Eds.), Findings of the Association for Computational Linguistics: NAACL 2022, Association for Computational Linguistics, Seattle, United States, 2022, pp. 1455–1468. doi:10.18653/v1/2022.findings-naacl.109. URL aclanthology.org/2022.findings-naacl.109/

[51] L. Liu, H. Jiang, P. He, W. Chen, X. Liu, J. Gao, J. Han, On the Variance of the Adaptive Learning Rate and Beyond, in: Proceedings of the Eighth International Conference on Learning Representations (ICLR 2020), 2020.

[52] T. Ridnik, E. Ben-Baruch, N. Zamir, A. Noy, I. Friedman, M. Protter, L. Zelnik-Manor, Asymmetric loss for multi-label classification, in: Proceedings of the IEEE/CVF International Conference on Computer Vision, 2021, pp. 82–91.

[53] T.-Y. Ross, G. Dolĺar, Focal loss for dense object detection, in: Proceedings of the IEEE Conference on Computer Vision and Pattern Recognition, 2017, pp. 2980–2988.

[54] C. Szegedy, V. Vanhoucke, S. Ioffe, J. Shlens, Z. Wojna, Rethinking the inception architecture for computer vision, in: Proceedings of the IEEE Conference on Computer Vision and Pattern Recognition, 2016, pp. 2818–2826.

[55] R. Müller, S. Kornblith, G. E. Hinton, When does label smoothing help?, Advances in Neural Information Processing systems 32 (2019).

[56] P. Khosla, P. Teterwak, C. Wang, A. Sarna, Y. Tian, P. Isola, A. Maschinot, C. Liu, D. Krishnan, Supervised Contrastive Learning, Advances in Neural Information Processing Systems 33 (2020) 18661–18673.

[57] N. Lin, G. Qin, G. Wang, D. Zhou, A. Yang, An effective deployment of contrastive learning in multi-label text classification, in: Findings of the Association for Computational Linguistics: ACL 2023, 2023, pp. 8730–8744.

[58] M. Pakdaman Naeini, G. Cooper, M. Hauskrecht, Obtaining Well Calibrated Probabilities Using Bayesian Binning, Proceedings of the AAAI Conference on Artificial Intelligence 29 (1) (Feb. 2015). doi:10.1609/aaai.v29i1.9602. URL ojs.aaai.org/index.php/AAAI/article/view/9602

[59] J. Bastings, K. Filippova, The elephant in the interpretability room: Why use attention as explanation when we have saliency methods?, in: Proceedings of the Third BlackboxNLP Workshop on Analyzing and Interpreting Neural Networks for NLP, 2020, pp. 149–155.

[60] M. Sundararajan, A. Taly, Q. Yan, Axiomatic attribution for deep networks, in: International Conference on Machine Learning, PMLR, 2017, pp. 3319–3328.

61. N. Kokhlikyan, V. Miglani, M. Martin, E. Wang, B. Alsallakh, J. Reynolds, A. Melnikov, N. Kliushkina, C. Araya, S. Yan, O. Reblitz-Richardson, Captum: A unified and generic model interpretability library for PyTorch (2020). arXiv:2009.07896.

[62] L. Hoang, Y. Guan, H. Kilicoglu, Methodological information extraction from randomized controlled trial publications: a pilot study, in: AMIA Annual Symposium Proceedings, Vol. 2022, 2022, p. 542.

[63] S. Tybaert. MEDLINE Data Changes—2019. NLM Tech Bull. 2018 Nov-Dec;(425):e4a.

[64] S. E. Doneva, S. de Viragh, H. Hubarava, S. Schandelmaier, M. Briel, B. V. Ineichen, Studytypeteller—large language models to automatically classify research study types for systematic reviews, Research Synthesis Methods (2025) 1–20.

[65] A. Lazaridou, A. Kuncoro, E. Gribovskaya, D. Agrawal, A. Liska, T. Terzi, M. Gimenez, C. de Masson d’Autume, T. Kocisky, S. Ruder, et al., Mind the gap: Assessing temporal generalization in neural language models, Advances in Neural Information Processing Systems 34 (2021) 29348–29363.

[66] E. Tutubalina, A. Kadurin, Z. Miftahutdinov, Fair evaluation in concept normalization: a large-scale comparative analysis for bert-based models, in: Proceedings of the 28th International Conference on Computational Linguistics, 2020, pp. 6710–6716.

[67] I. Alimova, E. Tutubalina, S. I. Nikolenko, Cross-domain limitations of neural models on biomedical relation classification, IEEE Access 10 (2021) 1432–1439.

[68] R. Islamaj, P.-T. Lai, C.-H. Wei, L. Luo, T. Almeida, R. A. A. Jonker, S. I. R. Conceicão, D. F. Sousa, C.-P. Phan, J.-H. Chiang, et al., The overview of the biored (biomedical relation extraction dataset) track at biocreative viii, Database 2024 (2024) baae069.

[69] Q. Chen, Y. Hu, X. Peng, Q. Xie, Q. Jin, A. Gilson, M. B. Singer, X. Ai, P.-T. Lai, Z. Wang, et al., Benchmarking large language models for biomedical natural language processing applications and recommendations, Nature communications 16 (1) (2025) 3280.

[70] M. Ma, G. Chochlakis, N. M. Pandiyan, J. Thomason, S. Narayanan, Large language models do multi-label classification differently, arXiv preprint arXiv:2505.17510 (2025).

[71] N. R. Smalheiser, D. P. Fragnito, E. E. Tirk, Anne O’Tate: Value-added PubMed search engine for analysis and text mining, PloS one 16 (3) (2021) e0248335.

